# Flatten the Curve! Modeling SARS-CoV-2/COVID-19 Growth in Germany at the County Level

**DOI:** 10.1101/2020.05.14.20101667

**Authors:** Thomas Wieland

**Affiliations:** Karlsruhe Institute of Technology

## Abstract

Since the emerging of the “novel coronavirus” SARS-CoV-2 and the corresponding respiratory disease COVID-19, the virus has spread all over the world. Being one of the most affected countries in Europe, in March 2020, Germany established several nonpharmaceutical interventions to contain the virus spread, including the closure of schools and child day care facilities (March 16-18, 2020) as well as a full “lockdown” with forced social distancing and closures of “nonessential” services (March 23, 2020). The present study attempts to analyze whether these governmental interventions had an impact on the declared aim of “flattening the curve”, referring to the epidemic curve of new infections. This analysis is conducted from a regional perspective. On the level of the 412 German counties, logistic growth models were estimated based on daily infections (estimated from reported cases), aiming at determining the regional growth rate of infections and the point of inflection where infection rates begin to decrease and the curve flattens. All German counties exceeded the peak of new infections between the beginning of March and the middle of April. In a large majority of German counties, the epidemic curve has flattened before the “lockdown” was established. In a minority of counties, the peak was already exceeded before school closures. The growth rates of infections vary spatially depending on the time the virus emerged. Counties belonging to states which established an additional curfew show no significant improvement with respect to growth rates and mortality. Furthermore, mortality varies strongly across German counties, which can be attributed to infections of people belonging to the “risk group”, especially residents of retirement homes. The results raise the question whether social ban measures and curfews really contribute to curve flattening within a pandemic.

## 1 Background

The “novel coronavirus” SARS-CoV-2 (“Severe Acute Respiratory Syndrome Coronavirus 2”) and the corresponding respiratory disease COVID-19 (“Coronavirus Disease 2019”) caused by the virus initially appeared in December 2019 in Wuhan, Province Hubei, China. Since its emergence, the virus has spread over nearly all countries across the world. On March 12, 2020, the World Health Organization (WHO) declared the SARS-CoV-2/COVID-19 outbreak a global pandemic (Lai et al. 2020, World Health Organization 2020b). As of May 10, 2020, 3,986,119 cases and 278,814 deaths had been reported worldwide. In Europe, the most affected countries are Spain, Italy, United Kingdom and Germany (European Centre for Disease Prevention and Control 2020).

The virus is transmitted between humans via droplets or through direct contact (Lai et al. 2020). In a very influential simulation study from March 2020, the Imperial College COVID-19 Response Team (Ferguson et al. 2020) suggested a series of public health measures aimed at slowing or stopping the transmission of the virus in absence of a vaccine or a successful therapy. These so-called *nonpharmaceutical interventions* (NPI) aim at reducing contact rates in the population, including social distancing and closures of schools and universities as well as the quarantine of infected persons. The Chinese government had imposed containment measures in the Provice Hubei already at the end of January 2020. This “lockdown” included a quarantine of the most affected city Wuhan and movement restrictions for the population as well as school closures (CNN 2020). In March 2020, nearly all European countries have introduced measures against the spread of Coronavirus. These measures range from appeals to voluntary behaviour changes in Sweden to strict curfews, e.g. in France and Spain (Deutsche Welle 2020a). The public health strategy to contain the virus spread is commonly known as “flatten the curve”, which refers to the epidemic curve of the number of infections: “Flattening the curve involves reducing the number of new COVID-19 cases from one day to the next. This helps prevent healthcare systems from becoming overwhelmed. When a country has fewer new COVID-19 cases emerging today than it did on a previous day, that’s a sign that the country is flattening the curve” (Johns Hopkins University 2020).

In Germany, due to the federal political system, measures to “flatten the curve” were introduced on the national as well as the state level. As the German “lockdown” has no single date, we distinguish here between four phases of NPIs, of which the main interventions were the closures of schools, child day care centers and most retail shops etc. in calendar week 12 (phase 2), and the nationwide establishment of a contact ban (attributed to phase 3), including forced social distancing and a ban of gatherings of all types, on March 23, 2020. The German states Bavaria, Saarland, and Saxony established additional curfews (see table 1). Occasionally, these governmental interventions were criticized because of the social, psychological and economic impacts of a “lockdown” and/or the lack of its necessity (Capital 2020, Süddeutsche Zeitung 2020a, Tagesspiegel 2020a, Welt online 2020a). Apart from the economic impacts emerging from a worldwide recession (The Guardian 2020), the psychosocial consequences of movement restrictions and social isolation (resulting from NPIs) have also become apparent now in terms of an increase of several mental health illnesses (Carvalho Aguiar Melo, de Sousa Soares 2020, Mucci et al. 2020, Williams et al. 2020). The effects of (forced) isolation as well as school and child day care closures are also visible through a worldwide increase in domestic abuse (New York Times 2020), reported in Germany as well (Stuttgarter Zeitung 2020, Süddeutsche Zeitung 2020b).

**Table 1:**
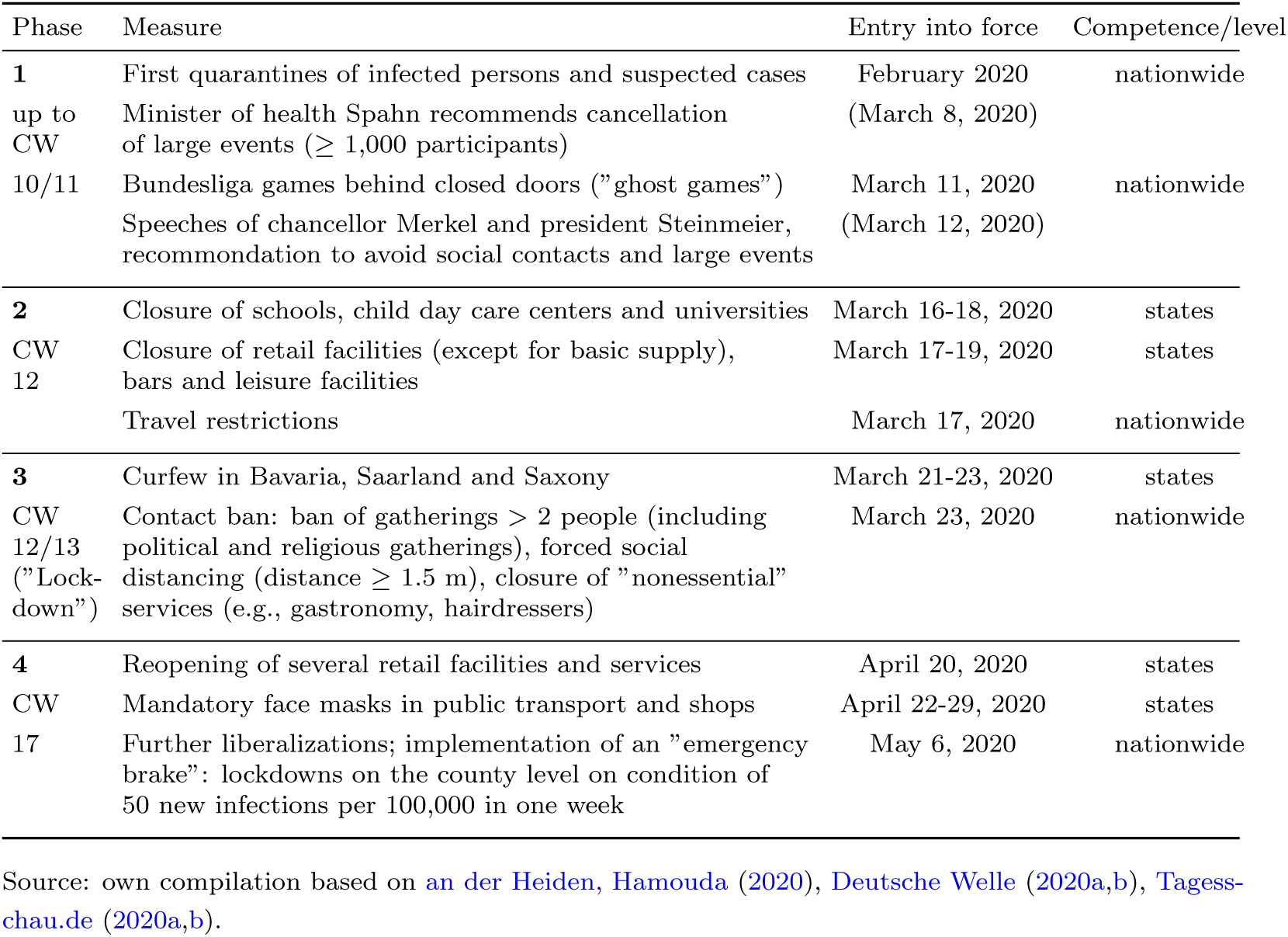
Main governmental nonpharmaceutical interventions with respect to COVID-19 pandemic in Germany

It is therefore all the more important to know whether these restrictions really contributed to the flattening of the epidemic curve of Coronavirus in Germany (RKI 2020a). This question should be addressed from a regional perspective for two reasons. *First*, in May 2020, the competences for the measures in Germany have shifted from the national to the state and regional (county) level. In the future, counties with more than 50 new infections per 100,000 in one week are expected to implement regional measures (see table 1). *Second*, a spatial perspective allows the impact of the German measures of March 2020 to be identified. In his statistical study, the mathematician Ben-Israel (2020) compares the epidemic curves of Israel, the USA and several European countries. These curves demonstrate a decline of new infections, regardless of the national measures to contain the virus spread. Furthermore, the study reveals the trend that the peak of infections is typically reached in the sixth week after the first case report, while a decline of the curve starts in the eighth week. This occurs in all assessed countries on the national level, no matter whether a “lockdown” was established (e.g. Italy) or not (e.g. Sweden).

The focus in the present study is on the main nonpharmaceutical interventions with respect to the SARS-CoV-2/COVID-19 pandemic in Germany, which means the concrete “lockdown” measures affecting the social and economic life of the whole society (distinguishing from measures taken in most cases of infectious dieseases, such as quarantine of affected persons). In the terminology of the present study, these are the phase 2 and 3 measures, denoted in table 1. Building upon the discrepancy outlined by Ben-Israel (2020), the present study addresses the following research questions:

- Pandemic or epidemic growth has a regional component due to regional infection hotspots or other behavorial or spatial factors. Thus, growth rates of infections may differ between regions in the same country (Chowell et al. 2014). In Germany, the prevalence of SARS-CoV-2/COVID-19 differs among the 16 German states and 412 counties, clearly showing “hotspots” in South German counties belonging to Baden-Wuerttemberg and Bavaria (RKI 2020a). Thus, the first question to be answered is: *How does the growth rate of SARS-CoV-2/COVID-19 vary across the 412 German counties?*
- The German measures to contain the pandemic entered into force nearly at the same time, especially in terms of closures of schools, childcare infrastructure and retailing (starting March 16/17, 2020) as well as the nationwide contact ban (starting March 23, 2020). Ben-Israel (2020) found a decline of new infection cases on the national level regardless of the Corona measures. To examine the effect of the German measures, we need to estimate the time of the peak and the declining of the curves of infection cases, respectively: *At which date(s) did the epidemic curves of SARS-CoV-2/COVID-19 in the 412 German counties flatten?*
- Regional prevalence and growth, as well as the mortality of SARS-CoV-2/COVID-19, are attributed within media discussions to several spatial factors, including population density or demographic structure of the regions (Welt online 2020b). Furthermore, the German measures differ on the state level, as three states - Bavaria, Saarland and Saxony - established additional curfews supplementing the other interventions (see table 1). Focusing on growth rate and mortality, and addressing these regional differences, the third research question is: *Which indicators explain the regional differences of SARS-CoV-2/COVID-19 growth rate and mortality on the level of the 412 German counties?*

## 2 Methodology

### 2.1 Logistic growth model

According to Li (2018), in simple terms, an infectious disease spread (pandemic or epidemic) can be summarized as follows: At the beginning, one or more infectious individuals are introduced into a population of *susceptibles* (non-infected/healthy individuals). As the pathogen (e.g., virus) is transmitted from one individual to another, the number of *infected* individuals increases over time. Depending on the regarded pathogen/disease, infected individuals *recover* due to medical interventions and/or reactions of the individuals’ immune system and, in many cases, gain partial or full immunity against the pathogen (e.g., through the development of antibodies against a virus). In other cases, infected people may also die from the disease. In all aforementioned cases and on condition of a stationary population, the number of susceptibles decreases and, thus, the number of new infections decreases as well. As a consequence, the pandemic/epidemic slows down and ends. The disease spread may also be contained by vaccination and/or other control and preventive measures. Note that, technically, one must distinguish between an *infection* and the *disease* which is (or may be) caused by the pathogen: “Disease is not the same as infection. Infection is said to have occurred when an organism successfully avoids innate defense mechanisms and stably colonizes a niche in the body. To establish an infection, the invader must first penetrate the anatomic and physiological barriers that guard the skin and mucosal surfaces of the host. Secondly, the organism must be able to survive in the host cellular milieu long enough to reproduce. This replication may or may not cause visible, clinical damage to the host tissues, symptoms that we call ‘disease’” (Mak, Saunders 2006).

Analyzing the transmission and spreading process of infectious diseases involves the utilization of mathematical models. Pandemic growth can be modeled by deterministic models such as the SIR (susceptible-infected-recovered) model and its extensions, or stochastic, phenomenological models such as the exponential or the logistic growth model. The former type of model does not depend on large empirical data on disease cases but requires additional information about the disease and the transmission process. The latter type of model is based on linear or nonlinear regression and only empirical data of infections and/or confirmed cases of disease (or death) is required for model estimation (Batista 2020a,b, Chowell et al. 2014, 2015, Li 2018, Ma 2020, Pell et al. 2018). Recently, there have already been several attempts to model the SARS-CoV-2 pandemic on the country (or even world) level, by using either the original or extended SIR model (Batista 2020b), the logistic growth model (Batista 2020a, Vasconcelos et al. 2020, Wu et al. 2020), or both (Zhou et al. 2020).

In the present case, we regard the spread of the Coronavirus primarily as an empirical phenomenon over space and time rather than its epidemiological characteristics. We therefore focus on 1) the regional growth speed of the pandemic and 2) the time when exponential growth ends and the infection rate decreases again. Apart from that, only infection cases and some further information are available, but not additional epidemiological information. Thus, the method of choice is a phenomenological regression model. In an early phase of an epidemic, when the number of infected individuals growths exponentially, an exponential function could be utilized for the phenomenological analysis (Ma 2020). However, officially reported SARS-CoV-2 infections in Germany (measured by the time of onset of symptoms) declined from mid-March and the corresponding reproduction number was estimated at *R* = 0.71 based on the case reports as of May 6, 2020 (RKI 2020a), which indicates that the phase of exponential growth was exceeded at this time. Thus, a logistic growth model is used for the analysis of SARS-CoV-2 growth in the German counties.

The following representations of the logistic growth model are adopted from Batista (2020a), Chowell et al. (2014) and Tsoularis (2002). Unlike exponential growth, logistic growth includes two stages, while assuming a saturation effect. The first stage is characterized by an exponential growth of infections due to an unregulated spreading of the disease. However, as more infections accumulate, the number of at-risk susceptible persons decreases because of immunization, death, or behavioral changes as well as public health interventions. After the inflection point of the infection curve, when the infection rate is at its maximum, the growth decreases and the cumulative number of infections approximate its theoretical maximum, which is the saturation value (see figure 1).

**Figure 1:**
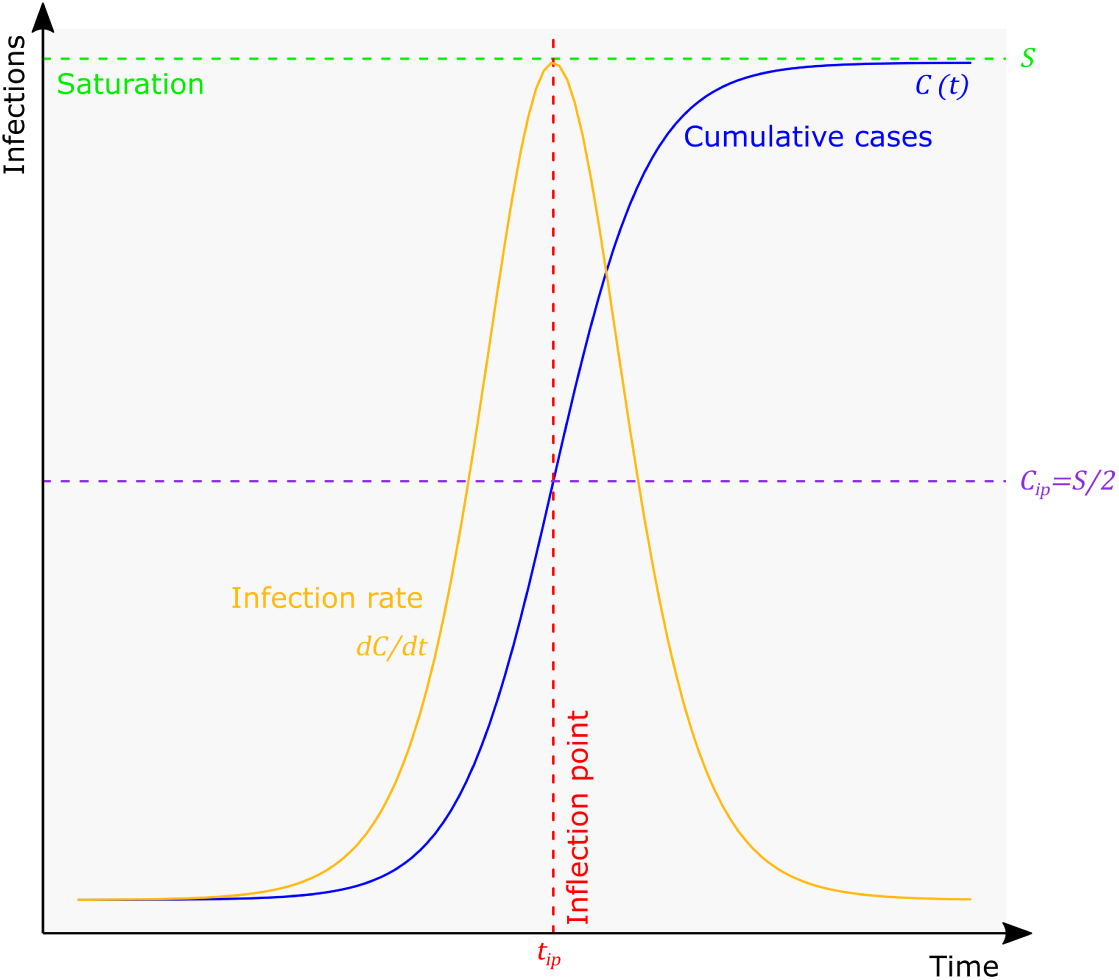
Logistic growth of an epidemic

In the logistic growth model, the cumulative number of infected or diseased persons at time *t, C*(*t*) is a function of time:

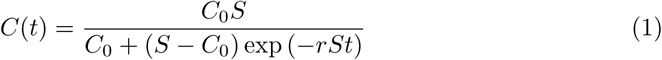

where *C*_0_ is the initial value of *C* at time 0, *r* is the intrinsic growth rate, and *S* is the saturation value.

The infection rate is the first derivative:

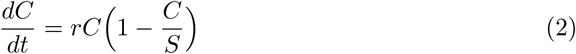

The inflection point of the logistic curve indicates the maximal infection rate before the growth declines, which means a flattening of the cumulative infection curve. The inflection point, *ip*, is equal to:

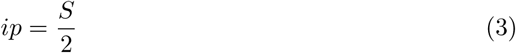

at time

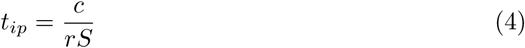

where:

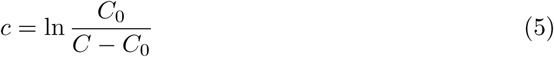

When empirical data (here: time series of cumulative infections) is available, the three model parameters *r, S* and *C*_0_ can be estimated empirically.

In the present case, fitting the models is done in a three-step estimation procedure including both OLS (Ordinary Least Squares) and NLS (Nonlinear Least Squares) estimation. The former is used for creating start values for the iterative NLS estimation, while making use of the linearization and stepwise parametrization of the logistic function. Following the representations of the linearization of the logistic growth model described in Engel (2010), the nonlinear logistic model (formula 1) can be transformed into a linear model (on condition that the saturation value is known) by taking the reciprocal on both sides, taking natural logarithms and rearranging the function:

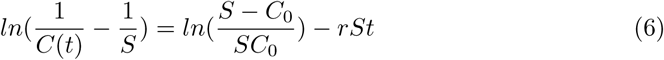

The transformed dependent variable, 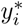, can be expressed by a linear relationship with two parameters, the intercept 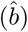 and slope 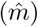:

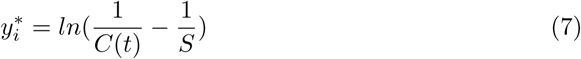

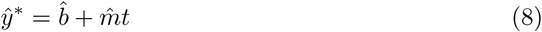

In step 1, an approximation of the saturation value is estimated, which is necessary for the linear transformation of the model. Transforming the empirical values *C*(*t*) according to formula 7, we have a linear regression model (formula 8). By utilizing bisection (Kaw et al. 2011), the best value for *S* is searched minimizing the sum of squared residuals. The bisection procedure consists of 10 iterations, while the start values are set around the current maximal value of *C*(*t*) (*max*(*C*(*t*)) + 1; *max*(*C*(*t*)) 1.2).

The resulting preliminary start value for saturation, *Ŝ*_*start*_, is used in step 2. We transform the observed *C*(*t*) using formula (7) with the preliminary value of *Ŝ* from step 1, *Ŝ*_*start*_. Another OLS model is estimated (formula 8). The estimated coefficients are used for calculating the start values of 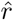 and *Ĉ*_0_ for the nonlinear estimation (Engel 2010):

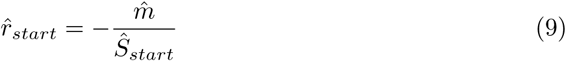

and

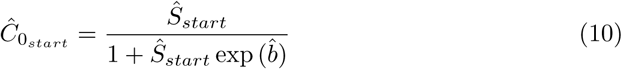

In step 3, the final model fitting is done using Nonlinear Least Squares (NLS), while inserting the values from steps 1 and 2, *Ŝ*_*start*_, 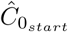 and 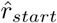, as start values for the iterative process. The NLS fitting uses default Gauss-Newton algorithm (Ritz, Streibig 2008) with a maximum of 500 iterations.

Using the estimated parameters 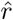, *Ĉ*_0_ and *Ŝ*, the inflection point of each curve is calculated via formulae 3 to 5. The inflection point *t*_*ip*_ is of unit time (here: days) and assigned to the respective date 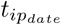 (YYYY-MM-DD). Based on this date, the following day 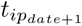 is the first day after the inflection point at which time the infection rate has decreased again. For graphical visualization, the infection rate is also computed using formula 2.

### 2.2 Estimating the dates of infection

In the present study, the daily updated data on confirmed SARS-CoV-2/COVID-19 cases, provided by federal authorities, the German Robert Koch Institute (RKI), is used (RKI 2020b). This dataset includes all persons which have been tested positive on the SARS-CoV-2 virus using a PCR (*polymerase chain reaction*) test and reported from local health authorities to the RKI. However, one must consider that neither the volume of tests nor the criteria for conducting a test are constant over time: Up to and including May 2020, almost exclusively people with acute respiratory symptoms were tested for SARS-CoV-2, as, with few exceptions, the presence of relevant symptoms is an exclusion criterion for testing in the RKI guidelines for medical doctors (IBBS 2020). In other words, this testing policy is targeted at the *disease* (COVID-19), not the *virus* (SARS-CoV-2). Thus, most of the cases in the present data are COVID-19 sufferers, whilst asymptomatic infected people and individuals with milder course are underrepresented. In the vast majority of cases, the date of onset of symptoms is reported in the dataset as well (an der Heiden, Hamouda 2020). The test volume was increased heavily from calendar week 11 (127,457 tests) to 12 (348,619 tests) but remains in the same order of magnitude until calendar week 18 (300,000-400,000 tests per week) (RKI 2020c).

The dataset used here is from May 5, 2020 and includes 163,798 cases. This data includes information about age group, sex, the related place of residence (county) and the date of report (Variable *Meldedatum*). The reference date in the dataset (Variable *Refdatum*) is either the day the disease started, which means the onset of symptoms, or the date of report (an der Heiden, Hamouda 2020). The date of onset of symptons is reported in the majority of cases (108,875 and 66.47 %, respectively).

The date of infection, which is of interest here, is either unknown or not included in the official dataset. Thus, it is necessary to estimate the approximate date of infection dependent on two time periods: the time between the infection and the onset of symptons (incubation period) and the delay between onset of symptoms and official report (reporting delay). Taking into account the 108,875 cases where the onset of symptoms is known, the mean delay between onset of symptons and case report is equal to 6.84 days. In the estimation, an incubation period equal to five days is assumed. This is a rather conservative assumption (which means a relatively short time period) referring to the current epidemiological estimates (see table 2). In their model-based scenario analysis towards the total number of diseases and deaths, the RKI also assumes an average incubation period of five days (an der Heiden, Buchholz 2020). Taking into account incubation period and reporting delay, there is an average all-over delay between infection and reporting of about 12 days (see figure 2).

**Table 2:** Studys estimating the incubation period of SARS-CoV-2/COVID-19

| Study | n | Distribution | Mean (CI95) | SD (CI95) | Median (CI95) | Min. | Max. |
| --- | --- | --- | --- | --- | --- | --- | --- |
| Backer et al. (2020) | 88 | Weibull | NA | 2.3 (1.7, 3.7) | 6.4 (5.6, 7.7) | NA | NA |
| Lauer et al. (2020) | 181 | Lognormal | 5.5 | 1.52 (1.3, 1.7) | 5.1 (4.5, 5.8) | NA | NA |
| Leung (2020) | 175 (a) | Weibull | 1.8 (1.0, 2.7) | NA | NA | NA | NA |
|  | 175 (b) | Weibull | 7.2 (6.1, 8.4) | NA | NA | NA | NA |
| Li et al. (2020) | 10 | Lognormal | 5.2 (4.1, 7.0) | NA | NA | NA | NA |
| Linton et al. (2020) | 158 | Lognormal | 5.6 (5.0, 6.3) | 2.8 (2.2, 3.6) | 5.0 (4.4, 5.6) | 2 | 14 |
| Sun et al. (2020) | 33 | NA | 4.5 | NA | NA | NA | NA |
| Xia et al. (2020) | 124 | Weibull | 4.9 (4.4, 5.4) | NA | NA | NA | NA |

Notes: (a) = Travelers to Hubei, (b) = Non-Travelers.
Source: own compilation.

**Figure 2:**
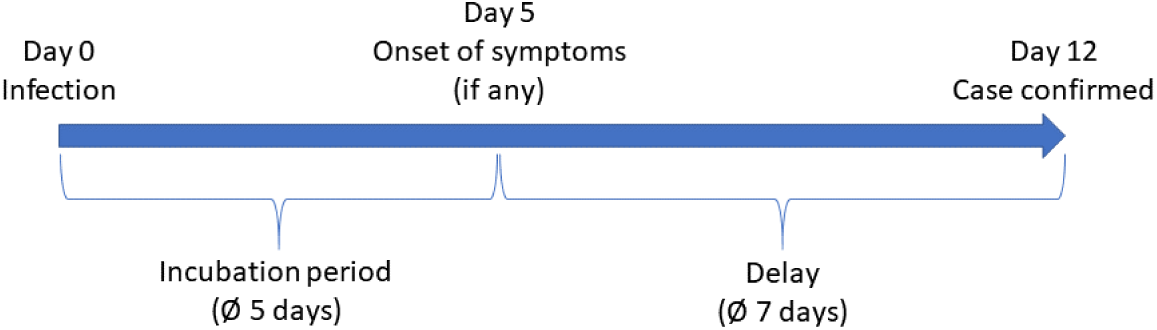
Time between infection and reporting of case

But this is just one side of the coin, as an inspection of the case data reveals that this delay differs between case characteristics (age group, sex) and counties. In their current prognosis, the RKI estimates the dates of onset of symptoms by Bayesian nowcasting based on the reporting date, but not taking into account the incubation time. The RKI nowcasting model incorporates delays of reporting depending on age group and sex, but not including spatial (county-specific) effects (an der Heiden, Hamouda 2020). Exploring the dataset used here, we see obvious differences in the reporting delay with respect to age groups and sex. There seems to be a tendency of lower reporting delays for young children and older infected individuals (see table 3). Taking a look at the delays between onset of symptoms and reporting date on the level of the 412 counties (not shown in table), the values range between 2.39 days (Würzburg city) and 17.0 days (Würzburg county).

**Table 3:** Delay between onset of symptoms and official report by age group and sex

| Age group | Sex | Delay between onset of symptoms<br>and reporting date [days] |  |
| --- | --- | --- | --- |
|  |  | Mean | SD |
| A00-A04 | female | 5.82 | 5.68 |
| A05-A14 | female | 6.09 | 5.34 |
| A15-A34 | female | 6.82 | 5.59 |
| A35-A59 | female | 7.00 | 5.98 |
| A60-A79 | female | 7.08 | 6.22 |
| A80+ | female | 5.10 | 5.83 |
| unknown | female | 8.71 | 9.79 |
| A00-A04 | male | 5.93 | 5.98 |
| A05-A14 | male | 6.04 | 5.12 |
| A15-A34 | male | 6.78 | 5.52 |
| A35-A59 | male | 7.18 | 5.90 |
| A60-A79 | male | 7.20 | 6.14 |
| A80+ | male | 5.70 | 5.84 |
| unknown | male | 9.86 | 8.42 |
| A00-A04 | unknown/diverse | 3.50 | 2.39 |
| A05-A14 | unknown/diverse | 4.00 | 5.20 |
| A15-A34 | unknown/diverse | 6.44 | 4.87 |
| A35-A59 | unknown/diverse | 6.95 | 5.51 |
| A60-A79 | unknown/diverse | 7.36 | 5.87 |
| A80+ | unknown/diverse | 6.50 | 11.40 |
| unknown | unknown/diverse | 9.60 | 3.58 |
| all-over |  | 6.84 | 5.90 |

Source: own calculation based on data from RKI \(2020b\).
Date of onset of symptoms is known for 108,875 (66.47 %) of 163,798 cases in the dataset.

For the estimation of the dates of infection, it is necessary to distinguish between the cases in which the date of symptom onset is known or not. In the former case, no assumption must be made towards the delay between onset of symptoms and date report. The calculation is simply:

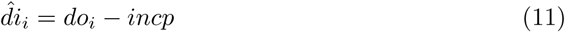

where 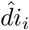 is the estimated date of infection of case *i, do*_*i*_ is the date of onset of symptoms reported in the RKI dataset and *incp* is the average incubation period equal to five (days).

For the 54,923 cases without information about onset of symptoms, we estimate this delay based on the 108,875 cases with known delays. As the reporting delay differs between age group, sex and county, the following dummy variable regression model is estimated (stochastic disturbance term is not shown):

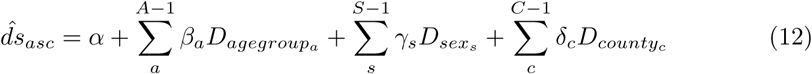

where 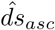 is the estimated delay between onset of symptoms and report depending on age group *a*, sex *s* and county *c*, 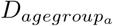 is a dummy variable indicating age group *a*, 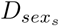 is a dummy variable indicating sex *s*, 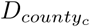is a dummy variable indicating county *c, A* is the number of age groups, *S* is the number of sex classifications, *C* is the number of counties and *α, β, γ* and *δ* are the regression coefficients to be estimated.

Taking into account the delay estimation, if the onset of symptoms is unknown, the date of infection of case *i* is estimated via:

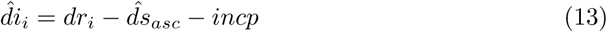

where *dr*_*i*_ is the date of report in the RKI dataset.

### 2.3 Models of regional growth rate and mortality

To test which variables predict the intrinsic growth rate and the regional mortality of SARS-CoV-2/COVID-19, respectively, two regression models were estimated. In the first model with the intrinsic growth rate *r* as dependent variable, we include the following predictors:

- In the media coverage about regional differences with respect to COVID-19 cases in Germany, several experts argue that a lower population density and a higher share of older population reduce the spread of the virus, with the latter effect being due to a lower average mobility (Welt online 2020b). It is well known that human mobility potentially increases the spread of an infectious disease. Also work-related commuting and tourism are considered as drivers of virus transmission (Charaudeau et al. 2014, Dalziel et al. 2014, Findlater, Bogoch 2018). To test these effects, four variables are included into the model: 1) The population density (*POPDENS*), 2) the share of population of at least 65 years (*POPS*65), 3) an indicator for the intensity of commuting (*CMI*) formulated by Guth et al. (2010), and 4) the number of annual tourist arrivals per capita (*TOUR*) for each county. All variables were calculated based on official statistics for the most recent year (2018/2019) (Destatis 2020a,b,c).
- In the media coverage, the lower prevalence in East Germany is also explained by 1) a different vaccination policy in the former German Democratic Republic and 2) a lower affinity towards carnival events as well as 3) less travelling to ski resorts due to lower incomes (Welt online 2020b). Thus, a dummy variable (1/0) for East Germany is included in the model (*EAST*).
- We test for the influence of different governmental interventions by including dummy variables for the states (“Länder”) Bavaria (*BV*), Saarland (*SL*), Saxony (*SX*) and North Rhine Westphalia (*NRW*), as well as Baden-Wuerttemberg (*BW*). Unlike the other 13 German states, the first three states established a curfew additional to the other measures at the time of phase 3, as is identified in the present study. Like Bavaria, North Rhine Westphalia and Baden-Wuerttemberg belong to the “hotspots” in Germany, with the latter state having a prevalence similar to Bavaria. Saxony has a prevalence below the national average (RKI 2020a).
- Apart from any interventions, when a disease spreads over time, also the susceptible population *must* decrease over time. As more and more individuals get infected (maybe causing temporal or lifelong immunization or, in other cases, death), there are continually fewer healthy people to get infected (Li 2018) (see also section 2.1). Consequently, regional growth speed *must* decrease with increasing regional prevalence and over time (and vice versa). Considering the specific case of SARS-CoV-2/COVID-19, as the outbreak differs within German counties (starting with “hotspots” like Heinsberg or Tirschenreuth county), we have to assume that differences in growth are due to different periods of time the virus is present and differences in the corresponding prevalence, respectively. Thus, two control variables are included in the model, the county-specific prevalence (*PRV*) and the number of days since the first (estimated) infection (*DAY S*).

In the second model for the explanation of regional mortality (*MRT*), five more independent variables have to be incorporated:

- From the epidemiological point of view, the “risk group” of COVID-19 for severe courses (and even deaths) is defined as people of 60 years and older. The arithmetic mean of deceased attributed to COVID-19 is equal to 81 years (median: 82 years). Out of 6,831 reported deaths on May 5 2020, 6,524 were of age 60 or older (95.51 %). This is, inter alia, because of outbreaks in residential homes for the elderly (RKI 2020a). Thus, the raw data from the RKI (RKI 2020b) was used to calculate the share of confirmed infected individuals of age 60 or older in all infected persons for each county (*INFS*60), which is included into the regression model for regional mortality.
- Several health-specific variables are found to influence the mortality risk (as well as the risk of severe course) of COVID-19. These individual-specific risk factors include, inter alia, diabetes, obesity, other respiratory diseases, or smoking (Engina et al. 2020, Selvan 2020). There are several possible health indicators which are unfortunately not available for German counties. Thus, the average health situation is captured by incorporating the average regional life expectancy into the model (*LEXP*), which is made available by the German Federal Institute for Research on Building, Urban Affairs and Spatial Development (BBSR) (BBSR 2020).
- On the regional level, air pollution was found to be a contributing factor to COVID-19 fatality (Ogen 2020, Wu et al. 2020). According to Wu et al. (2020), the regional air pollution with respect to particulate matter (annual mean of daily *PM*_10_ values, unit: *μg/m*^3^) is included into the model (*PM*10). As Ogen (2020) shows a correlation between nitrogen dioxide concentration and COVID-19 fatality, this type of air pollution (annual mean of daily *NO*_2_ values, unit: *μg/m*^3^) is incorporated into the model as well (*NO*2). Both air quality indicators are made available by the German Environment Agency (UBA 2020a) on the level of single monitoring stations. These stations are available geocoded (UBA 2020b) and have been assigned to the German counties via a nearest neighbor join. Thus, the county-level values of both indicators equal the values of the nearest monitoring station.
- The intrinsic growth rate of each county is incorporated into the model as well. Considering the chronology of an infectious disease spread, there must be a reciprocal relationship between growth speed and mortality: The more individuals die in the context of the regarded disease, the less susceptibles are left to be infected, resulting in a deceleration of the pandemic spread (Li 2018) (see also section 2.1). Thus, there *must* be a negative correlation between mortality and growth rate, all other things being equal. Consequently, the county-specific intrinsic growth rate (*r*) is included as control variable.

See table 4 for all variables included into the models. All continuous variables, including the dependent variables (*r* and *MRT*, respectively), were transformed via natural logarithm in the regression analysis. This leads to an interpretation of the regression coefficients in terms of elasticities and semi-elasticities (Greene 2012). Two variants were estimated for the growth rate model (with and without dummy variables) and three for the mortality model (with and without growth rate as well as a third model including both growth rate and dummy variables). The minimum significance level was set to *p* ≤ 0.1. In the first step, the regression models were estimated using an Ordinary Least Squares (OLS) approach and tested with respect to multicollinearity using variance inflation factors (*V IF*) with a critical value equal to five (Greene 2012).

**Table 4:**
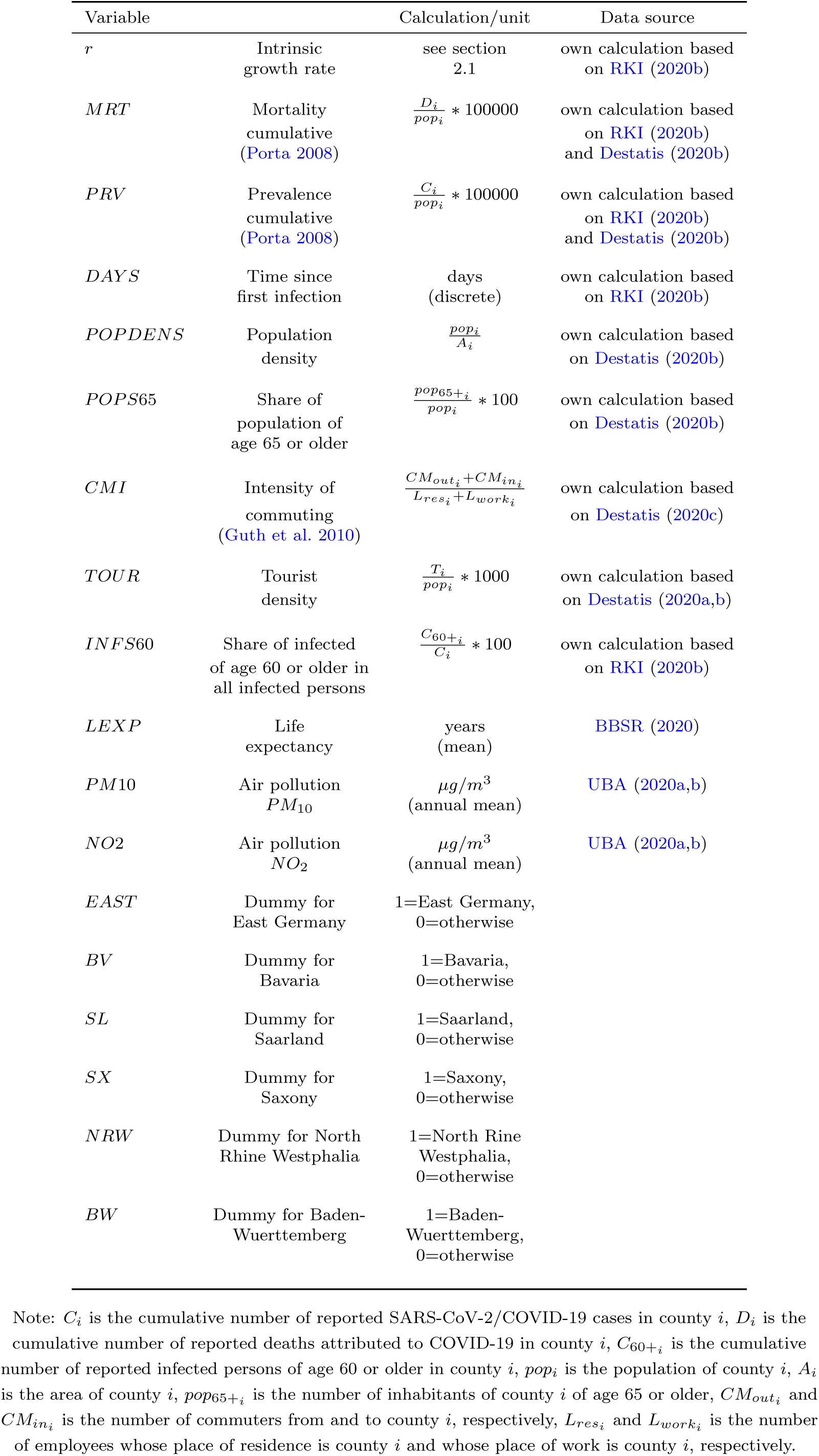
Variables in the regression models for growth rate and mortality

However, SARS-CoV-2/COVID-19 cases are obviously not evenly distributed across all German counties as the disease spread started in a few “hotspots” in Bavaria, Baden-Wuerttemberg and North Rhine Westphalia (RKI 2020a, Tagesspiegel 2020b). Of course, an infectious disease can be transmitted across county borders, in particular, by contact between residents of one region and a nearby region. As a consequence, it is to be expected that indicators of disease spread - such as the regarded variables growth rate and mortality - are similar between nearby regions. Thus, further model-based analyses require considering possible spatial autocorrelation in the dependent variables (Griffith 2009). Consequently, both dependent variables were tested for spatial autocorrelation using Moran’s I-statistic and the model estimation was repeated using a spatial lag model. In this type of regression model, spatial autocorrelation is modeled by a linear relationship between the dependent variable and the associated spatially lagged variable, which is a spatially weighted average value of the nearby objects. The influence of spatial autocorrelation is captured by adding a further parameter, *ρ* (*Rho*), to the regression equation, which is also tested for significance. Spatial linear regression models are not fitted by OLS but by Maximum Likelihood (ML) estimation. Both Moran’s I and the spatial lag model require a weighting matrix to define the proximity of the regarded spatial object to the other nearby objects (Chi, Zhu 2008, Rusche 2008). Here, the weighting matrix for the spatial object (county *i*) was defined as all adjacent counties.

### 2.4 Software

The analyses in this study was executed in `R` (R Core Team 2019), version 3.6.2. The parametrization of logistic growth models was done using own functions for the OLS estimation based on the description in Engel (2010) and the `nls()` function for the final NLS estimation. For the steps of the regression analyses and presentation of results, the packages `car` (Fox, Weisberg 2019), `REAT` (Wieland 2019), `spdep` (Bivand et al. 2013), and `stargazer` (Hlavac 2018) were used. For creating maps, `QGIS` (QGIS Development Team 2019), version 3.8, was used, including the plugin `NNJoin` (Tveite 2019) for one further analysis.

## 3 Results

### 3.1 Estimation of infection dates and national inflection point

Figure 3 shows the estimated dates of infection and dates of report of confirmed cases and deaths for Germany. The curves are not shifted exactly by the average delay period because of the different delay times with respect to case characteristics and county. The average time interval between estimated infection and case reporting is 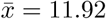 [days] (*SD* = 5.21). When applying the logistic growth model to the estimated dates of infection in Germany, the inflection point for whole Germany is at March 20, 2020.

**Figure 3:**
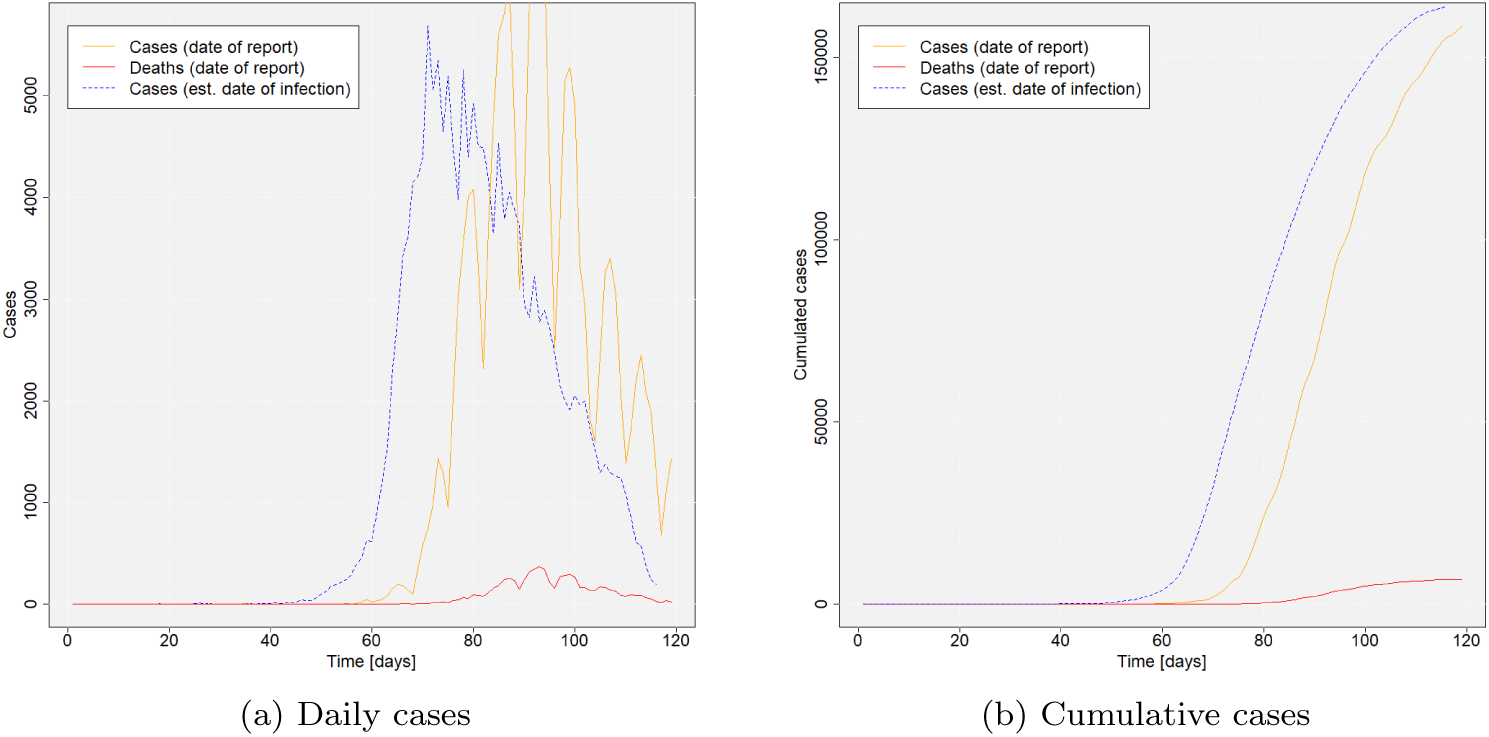
Reported SARS-CoV-2 infections in Germany over time (dates of report vs. estimated dates of infection) Source: own illustration. Data source: own calculations based on RKI (2020b)

Before switching to the regional level, we take into account the statistical uncertainty with respect to the estimation of the infection dates. About one third of the delay values for the time between onset of symptoms and case reporting was estimated by a stochastic model. Furthermore, the estimates of SARS-CoV-2/COVID-19 incubation period differ from study to study. This is why in the present case a conservative - which means a small - value of five days was assumed. Thus, we compare the results when including 1) the 95% confidence intervals of the response from the model in formula 12, and 2) the 95% confidence intervals of the incubation period as estimated by Linton et al. (2020). Figure 4 shows three different modeling scenarios, the mean estimation and the lower and upper bound of incubation period and delay time, respectively. The lower bound variant incorporates the lower bound of both incubation period and delay time, resulting in smaller delay between infection and case reporting and, thus, a later inflection point. The upper bound shows the counterpart. On the basis of the upper bound, the inflection point is already on March 19. Using the higher values of incubation period estimated by Backer et al. (2020), the upper bound results in an inflection point at March 17, while the lower bound variant leads to the turn on March 20. Considering confidence intervals of incubation period and delay time, the inflection point for the whole of Germany can be determined as occurring between March 17 and March 20, 2020 (see table 5).

**Table 5:** Date of inflection point depending on assumed incubation period

| Study | Median of incubation<br>time (CI-95) | Date of inflection point |  |  |
| --- | --- | --- | --- | --- |
|  |  | Lower | Median | Upper |
| Linton et al. (2020) | 5.0 (4.4, 5.6) | 2020-03-20 | 2020-03-20 | 2020-03-19 |
| Backer et al. (2020) | 6.4 (5.6, 7.7) | 2020-03-20 | 2020-03-19 | 2020-03-17 |

Source: own calculation based on data from RKI \(2020b\).

**Figure 4:**
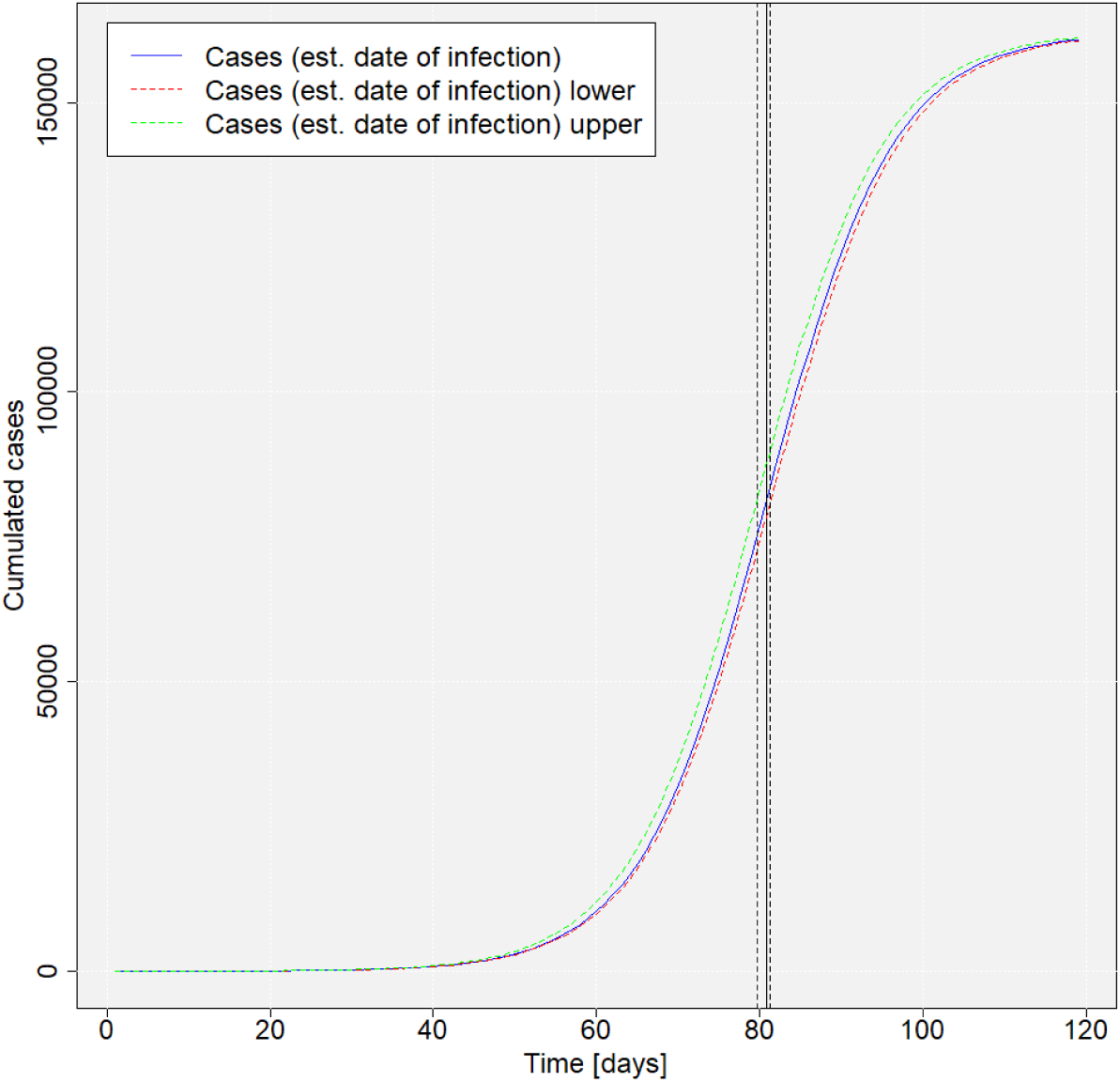
Estimated logistic growth model (including inflection point) for cumulative SARS-CoV-2 infections in Germany (based on estimated dates of infection) incorporating upper and lower bounds (95%-CI) Source: own illustration. Data source: own calculations based on RKI (2020b)

### 3.2 Estimation of growth rates and inflection points on the county level

Figure 5 shows the estimated intrinsic growth rates (*r*) for the 412 counties. Figure 6 provides six examples of the logistic growth curves with respect to four counties identified as “hotspots” (Tirschenreuth, Heinsberg, Greiz and Rosenheim) and two counties with a low prevalence (Flensburg and Uckermark). There are obviously differences in the growth rates, following a spatial trend: The highest growth rates can be found in counties in North Germany (especially Lower Saxony and Schleswig-Holstein) and East Germany (especially Mecklenburg-Western Pomerania, Thuringia and Saxony). To the contrary, the growth rates in Baden-Wuerttemberg and North Rine Westphalia appear to be quite low. Taking a look at the time the disease is present in the German counties, measured by the first estimated infection date (see figure 7), the growth rates tend be much smaller the longer the time since the disease appeared in the county.

**Figure 5:**
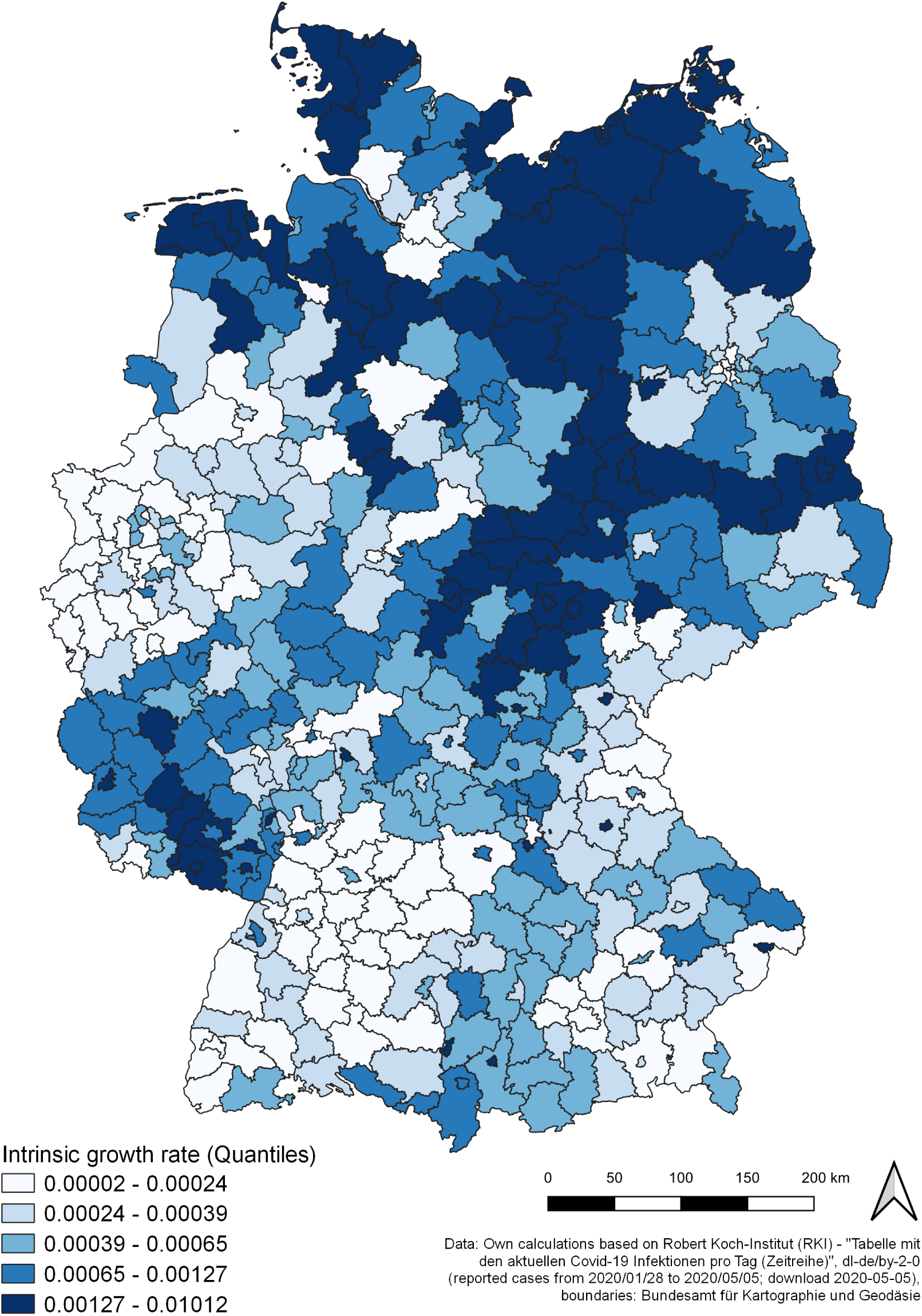
Intrinsic growth rate by county

**Figure 6:**
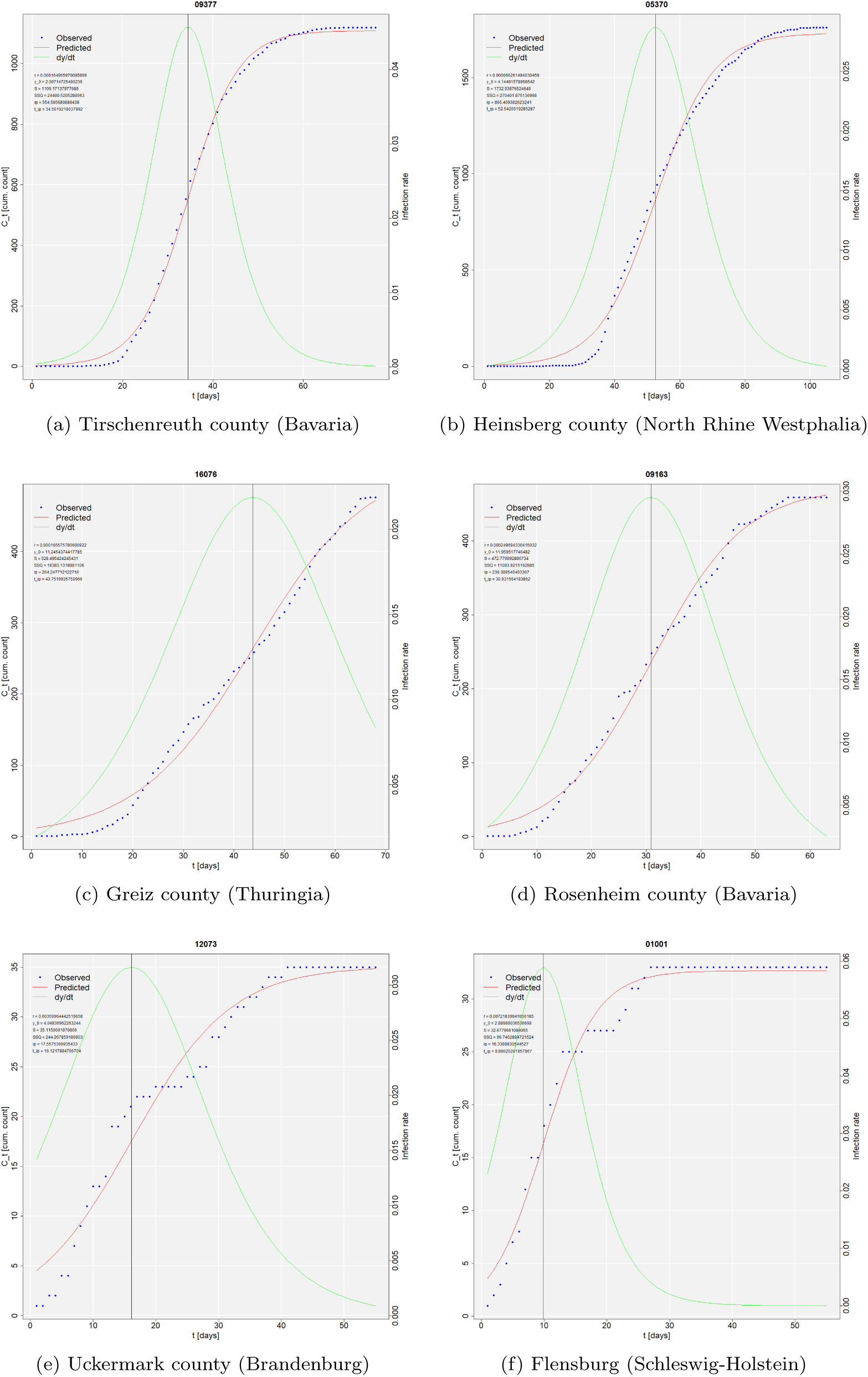
Cumulative SARS-CoV-2 infections (based on estimated dates of infection) and estimated logistic growth models (including infection rate and inflection point) in six German counties Source: own illustration. Data source: own calculations based on RKI (2020b)

**Figure 7:**
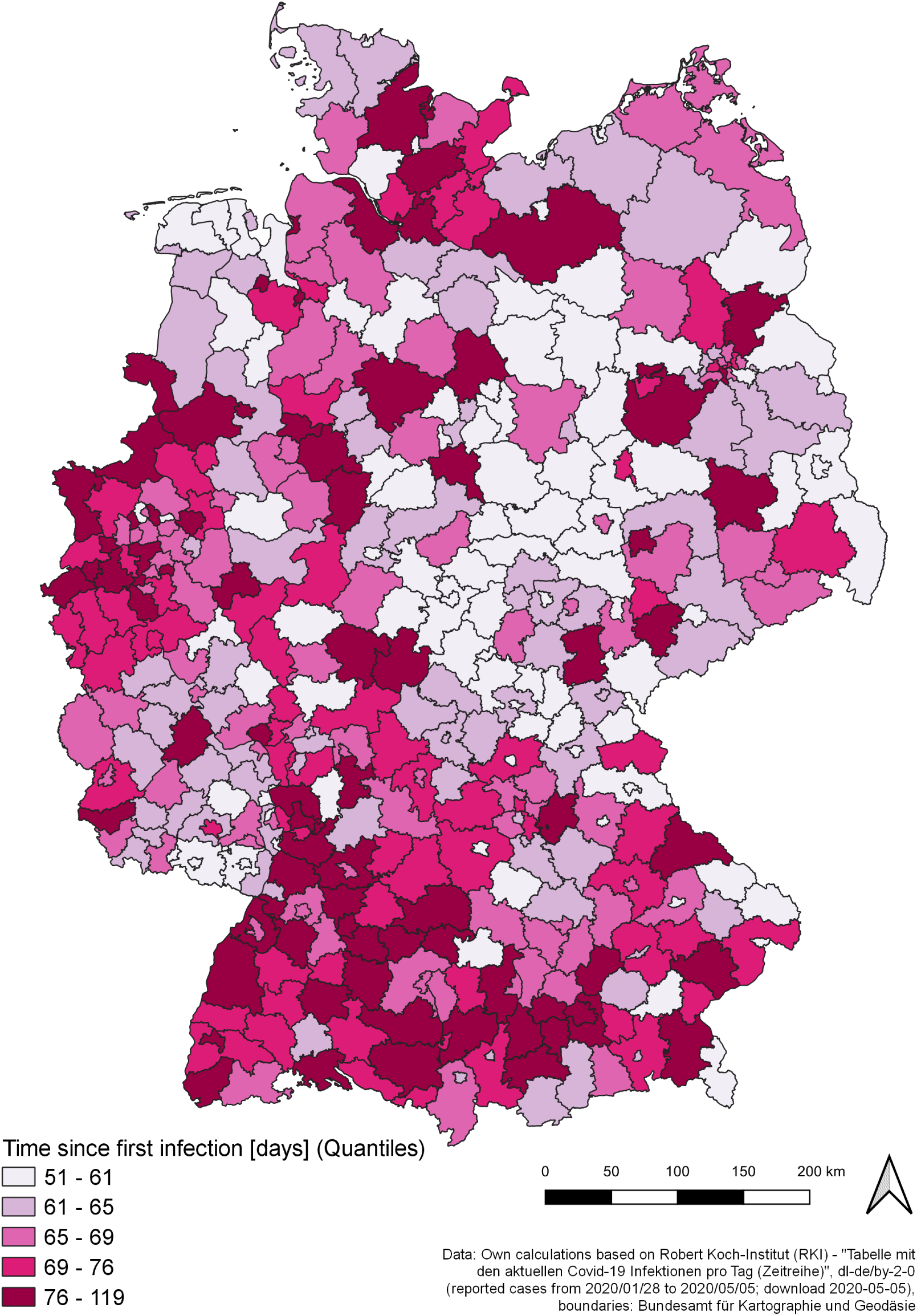
Time since first infection by county Source: own illustration.

The regional inflection points indicate the day with the local maximum of infection rate. From this day forth, the exponential disease growth turns into degressive growth. In figure 8, the dates of the first day after the regional inflection point are displayed, while categorizing the dates according to the coming into force of relevant nonpharmaceutical interventions (see table 1). Table 6 summarizes the number of counties and the corresponding population shares by these categories. Figure 9 shows the intrinsic growth rate (*y* axis) and the day after the inflection point (colored points) against time (*x* axis). Figure 10 shows the same information against regional prevalence (*x* axis).

**Table 6:** German counties by first day after inflection point

| First day after inflection point | Counties [no.] | Counties [%] | Population [Mill.] | Population [%] |
| --- | --- | --- | --- | --- |
| Before March 13 | 6 | 1.46 | 0.98 | 1.17 |
| March 13 to March 16 | 45 | 10.92 | 8.27 | 9.95 |
| March 17 to March 20 | 138 | 33.50 | 31.03 | 37.33 |
| March 21 to March 22 | 66 | 16.02 | 14.30 | 17.20 |
| March 23 to April 19 | 157 | 38.11 | 28.54 | 34.34 |
| Sum | 412 | 100 | 83.13 | 100 |

Source: own illustration. Data source: own calculations based on RKI \(2020b\)

**Figure 8:**
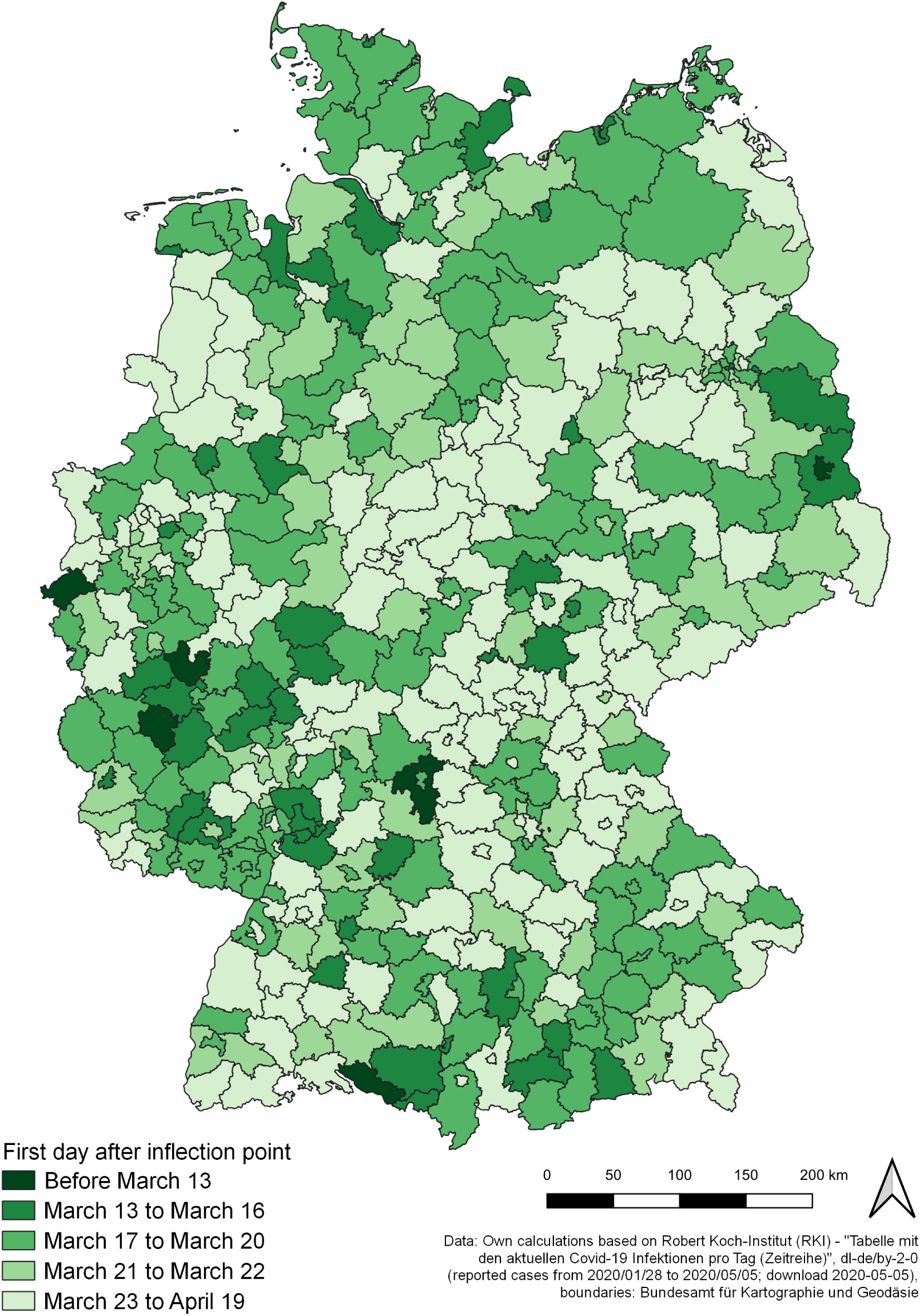
First day after inflection point by county Source: own illustration.

**Figure 9:**
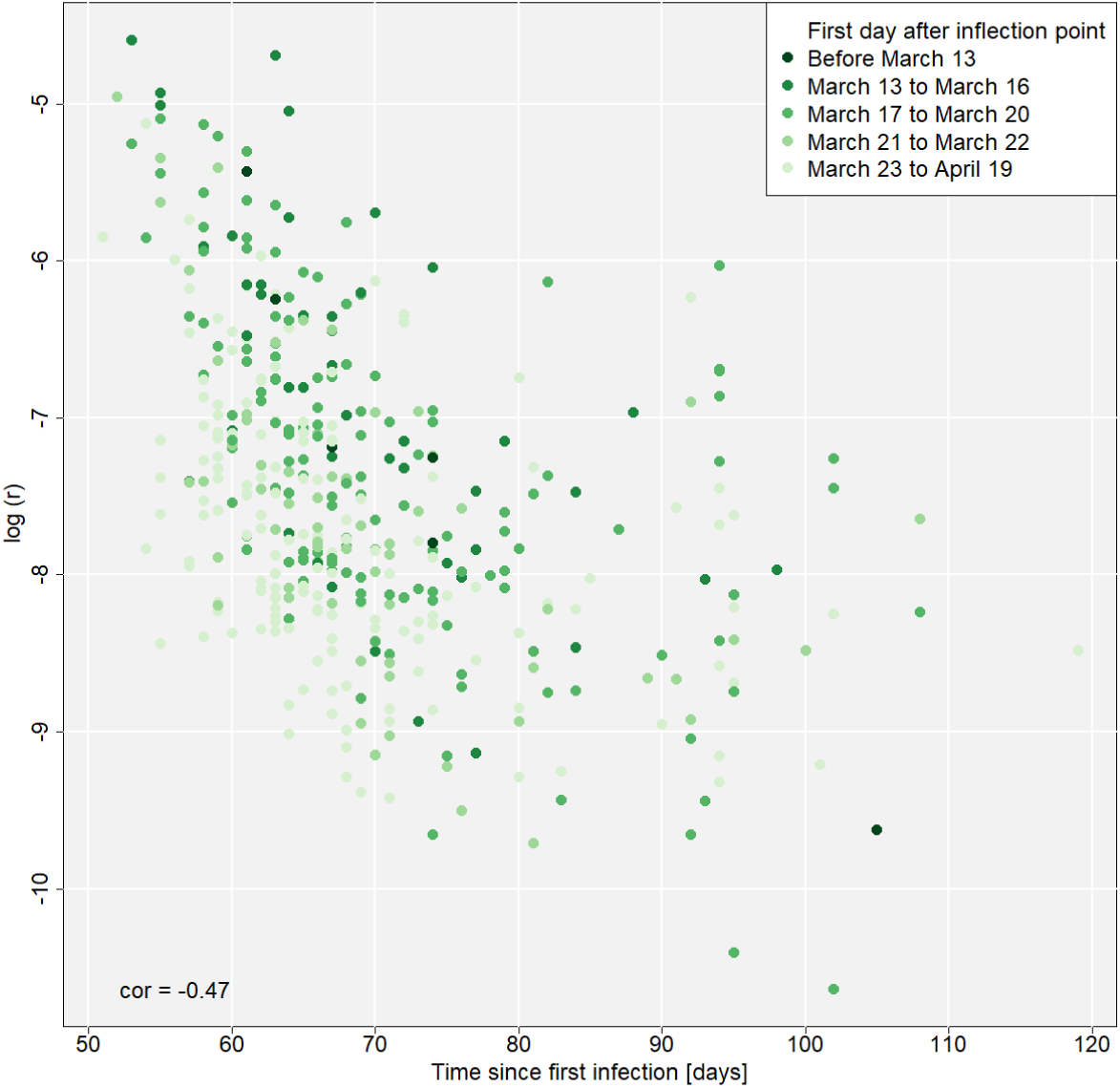
Growth rate and first day after inflection point vs. time Source: own illustration. Data source: own calculations based on RKI (2020b)

**Figure 10:**
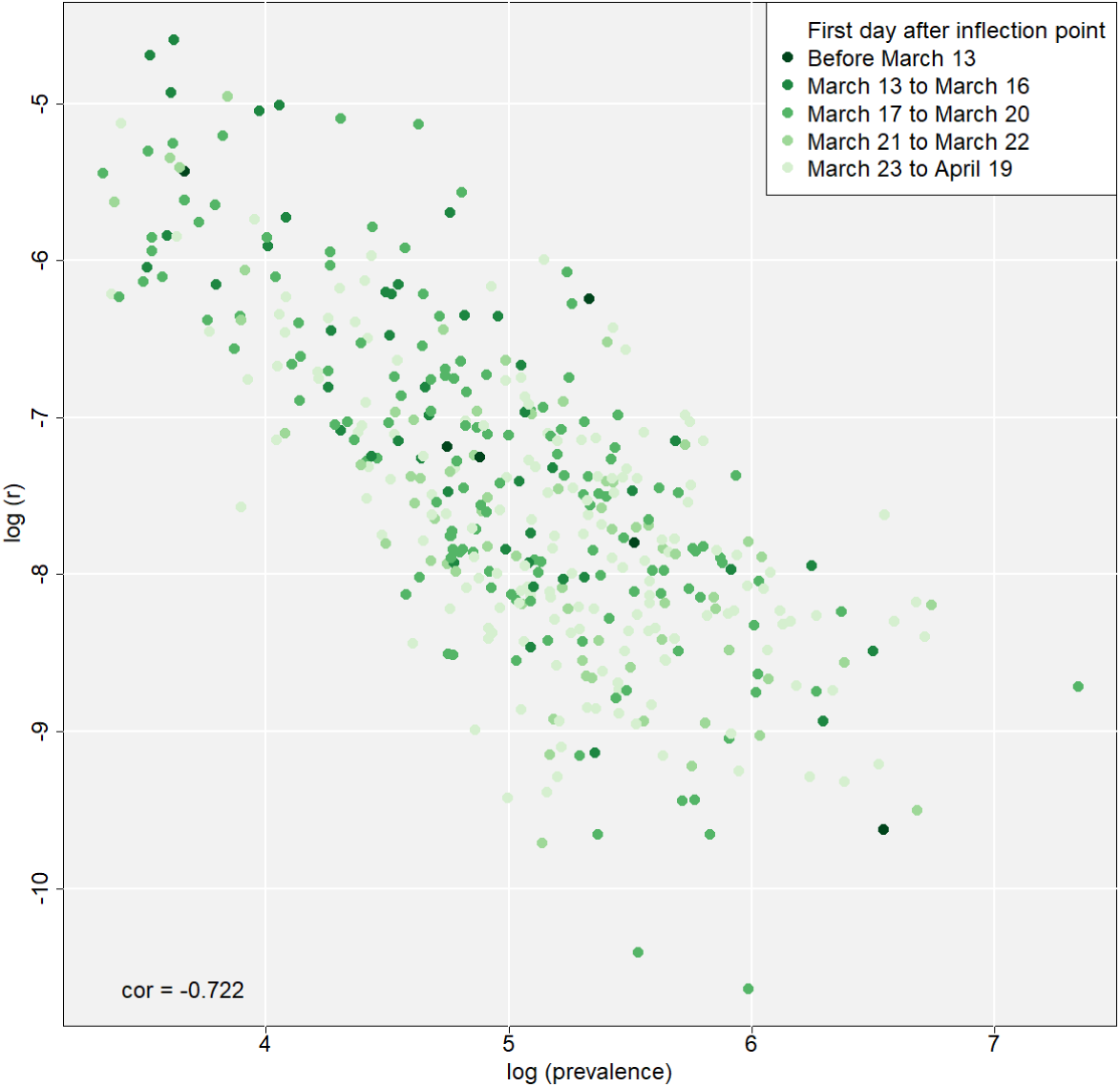
Growth rate and first day after inflection point vs. prevalence Source: own illustration. Data source: own calculations based on RKI (2020b)

In 255 of 412 counties (61.89 %) with 54.58 million inhabitants (65.66 % of the national population), the SARS-CoV-2/COVID-19 infections had already decreased before forced social distancing etc. (phase 3 of measures) came into force (March 23, 2020). In a minority of counties (51, 12.38 %), the curve already flattened before the closing of schools and child day care centers (March 16-18, 2020), six of them exceeded the peak of new infections even before March 13 (This category refers to the appeals of chancellor Merkel and president Steinmeier on March 12). In 157 counties (38.11 %) with a population of 28.54 million people (34.34 % of the national population), the decrease of infections took place within the time of strict regulations towards social distancing and ban of gatherings.

The average time interval between the first estimated infection and the respective inflection point of the county is 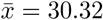 [days]. However, the time until inflection point is characterized by a large variance (*SD* = 11.92), but this may be explained partially by the (de facto unknown) variance in the incubation period and the variance in the delay between onset of symptoms and reporting date.

In all counties, the inflection points lie within March 6 and April 18, 2020, which means a time period of 43 days between the first and the last flattening of a county’s epidemic curve. The first decrease can be determined in Heinsberg county (North Rhine Westphalia; 254,322 inhabitants), which was one of the first Corona “hotspots” in Germany. The estimated inflection point here took place at March 6, 2020, leading to a date of the first day after the inflection point of March 7. The latest estimated inflection point (April 18, 2020) took place in Steinburg county (Schleswig-Holstein; 131,347 inhabitants).

As figure 9 shows, the intrinsic growth rates, which indicate an average growth level over time, and inflection points of the logistic models are linked to each other. Growth speed declines over time (see also the maps in figures 5 and 9), more precisely, it declines in line with the time the disease is present in the regarded county (Pearson correlation coefficient of −0.47, *p* < 0.001). The longer the time between inflection point and now, the lower the growth speed, and vice versa. This process takes place over all German counties with a time delay depending on the first occurrence of the disease. As shown in figure 10, there is also a negative correlation between regional prevalence and growth rate (Pearson correlation coefficient of −0.722, *p* < 0.001). These relationships, which are closely linked to the chronology of an infectious disease spread and the characteristics of the logistic growth model, respectively, are included into the regression models as control variables.

### 3.3 Regression models for intrinsic growth rates and mortality

The variables of most interest used within the models are displayed in the maps in figures 11 (prevalence, *PRV*), 12 (mortality, *MRT*) and 13 (share of infected individuals of age ≥ 60, *POPS*65). Additionally, figure 14 shows the current case fatality rate on the county level. Tables 7 and 8 show the estimation results for the OLS regression models explaining the intrinsic growth rates and the mortality, respectively, both transformed via natural logarithm. Table 9 displays the Moran’s I-statistic for the dependent variables of the two models. Tables 10 and 11 show the estimation results for the spatial lag models.

**Table 7:** Estimation results for the growth rate model (OLS)

|  | *Dependent variable:* |  |
| --- | --- | --- |
|  | ln (r) |  |
|  | (1) | (2) |
| ln (POPDENS) | −0.177***<br>(0.028) | −0.102***<br>(0.027) |
| ln (POPS65) | 0.830***<br>(0.284) | 1.165***<br>(0.269) |
| ln (CMI) | 0.607***<br>(0.091) | 0.420***<br>(0.087) |
| ln (TOUR) | 0.146***<br>(0.042) | 0.035<br>(0.041) |
| EAST | −0.087<br>(0.091) | −0.045<br>(0.089) |
| BV |  | 0.473***<br>(0.091) |
| SL |  | 0.118<br>(0.231) |
| SX |  | −0.580***<br>(0.168) |
| NRW |  | −0.428***<br>(0.097) |
| BW |  | 0.011<br>(0.108) |
| ln (PRV) | −0.911***<br>(0.049) | −1.039***<br>(0.056) |
| DAYS | −0.019***<br>(0.003) | −0.014***<br>(0.003) |
| Constant | −3.660***<br>(1.144) | −4.214***<br>(1.079) |
| Observations | 412 | 412 |
| R<sup>2</sup> | 0.678 | 0.731 |
| Adjusted R<sup>2</sup> | 0.672 | 0.723 |
| Residual Std. Error | 0.587 (df = 404) | 0.540 (df = 399) |
| F Statistic | 121.278*** (df = 7; 404) | 90.307*** (df = 12; 399) |
| *Note:* *p<0.1; **p<0.05; ***p<0.01 |  |  |

**Table 8:** Estimation results for the mortality model (OLS)

|  | *Dependent variable:* |  |  |
| --- | --- | --- | --- |
|  | ln (MRT+0.0001) |  |  |
|  | (1) | (2) | (3) |
| ln (POPDENS) | −0.060<br>(0.132) | −0.268**<br>(0.121) | −0.148<br>(0.124) |
| ln (POPS65) | −0.738<br>(1.316) | 0.883<br>(1.196) | 1.730<br>(1.230) |
| ln (CMI) | −0.627<br>(0.412) | 0.397<br>(0.385) | 0.025<br>(0.396) |
| ln (TOUR) | −0.265<br>(0.188) | 0.066<br>(0.173) | −0.081<br>(0.178) |
| ln (LEXP) | 54.205***<br>(12.259) | 11.409<br>(11.877) | 12.349<br>(12.556) |
| ln (PM10) | 0.053<br>(0.716) | −0.226<br>(0.645) | −0.200<br>(0.653) |
| ln (NO2) | 0.428<br>(0.287) | 0.178<br>(0.259) | 0.213<br>(0.259) |
| ln (INFS60+0.0001) | 0.618***<br>(0.104) | 0.459***<br>(0.095) | 0.462***<br>(0.094) |
| EAST | −0.980**<br>(0.395) | −0.496<br>(0.359) | −0.148<br>(0.385) |
| BV |  |  | 1.110***<br>(0.334) |
| SL |  |  | 0.800<br>(0.983) |
| SX |  |  | −0.909<br>(0.738) |
| NRW |  |  | −0.144<br>(0.433) |
| BW |  |  | 0.193<br>(0.465) |
| DAYS | 0.017<br>(0.013) | −0.014<br>(0.012) | −0.011<br>(0.012) |
| ln (r) |  | −1.570***<br>(0.161) | −1.535***<br>(0.171) |
| Constant | −237.384***<br>(55.406) | −62.529<br>(53.006) | −69.684<br>(55.923) |
| Observations | 412 | 412 | 412 |
| R<sup>2</sup> | 0.238 | 0.384 | 0.407 |
| Adjusted R<sup>2</sup> | 0.219 | 0.367 | 0.383 |
| Residual Std. Error | 2.596 (df = 401) | 2.336 (df = 400) | 2.308 (df = 395) |
| F Statistic | 12.525*** (df = 10; 401) | 22.679*** (df = 11; 400) | 16.916*** (df = 16; 395) |

Note:
\*p<0.1; \*\*p<0.05; \*\*\*p<0.01

**Table 9:** Moran’s I-statistic for intrinsic growth rates and mortality (Weighting matrix: all adjacent counties)

| Indicator | Moran’s I | Expectation | Variance | Standard deviate |
| --- | --- | --- | --- | --- |
| $\ln(r)$ | 0.4856*** | -0.0024 | 0.0011 | 14.738 |
| $\ln(MRT+0.0001)$ | 0.1635*** | -0.0024 | 0.0011 | 5.0555 |
| *Note:* |  |  | *p<0.1; **p<0.05; ***p<0.01 |  |

Source: own calculation.

**Table 10:**
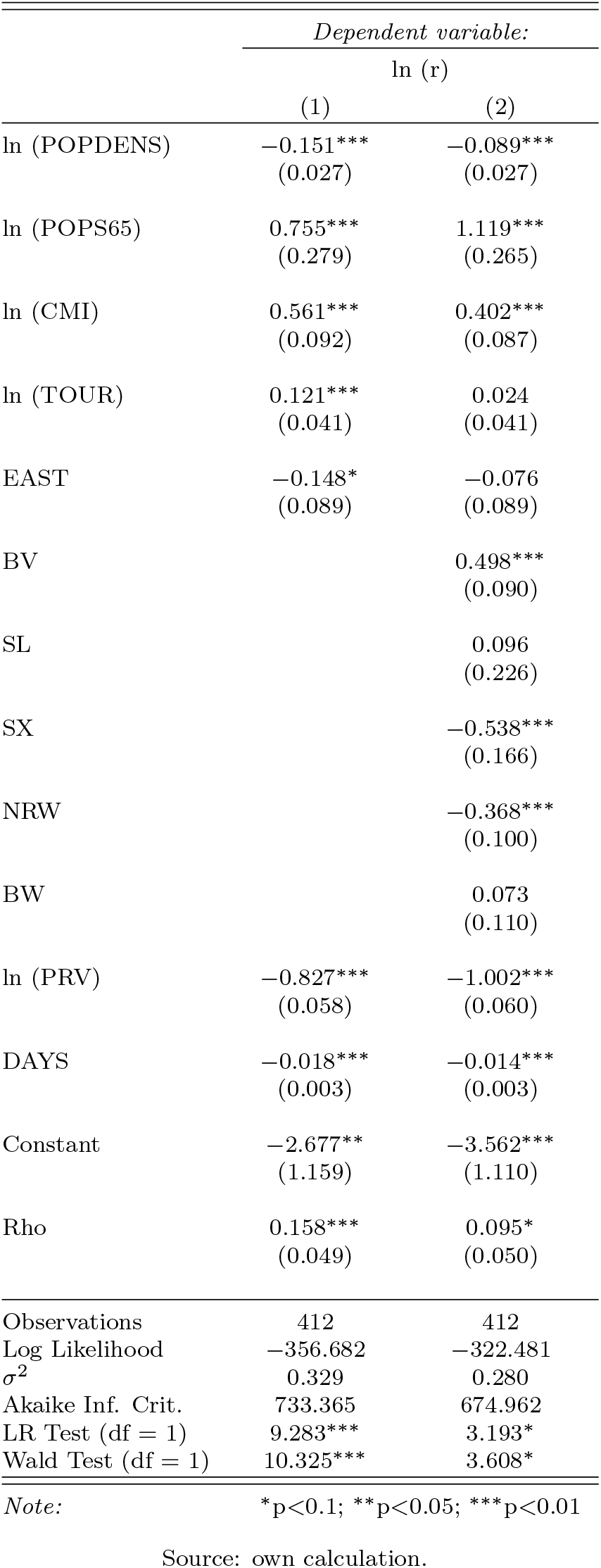
Estimation results for the growth rate model (spatial lag model)

**Table 11:** Estimation results for the mortality model (spatial lag model)

|  | *Dependent variable:* |  |  |
| --- | --- | --- | --- |
|  | ln (MRT+0.0001) |  |  |
|  | (1) | (2) | (3) |
| ln (POPDENS) | −0.085<br>(0.129) | −0.272**<br>(0.119) | −0.138<br>(0.121) |
| ln (POPS65) | −0.500<br>(1.289) | 0.921<br>(1.178) | 1.731<br>(1.204) |
| ln (CMI) | −0.586<br>(0.403) | 0.394<br>(0.379) | 0.013<br>(0.387) |
| ln (TOUR) | −0.250<br>(0.184) | 0.066<br>(0.170) | −0.086<br>(0.175) |
| ln (LEXP) | 51.311***<br>(12.101) | 11.262<br>(11.713) | 12.321<br>(12.288) |
| ln (PM10) | 0.197<br>(0.700) | −0.186<br>(0.637) | −0.224<br>(0.639) |
| ln (NO2) | 0.397<br>(0.280) | 0.174<br>(0.255) | 0.215<br>(0.254) |
| ln (INFS60+0.0001) | 0.614***<br>(0.102) | 0.460***<br>(0.094) | 0.461***<br>(0.092) |
| EAST | −0.764*<br>(0.399) | −0.448<br>(0.365) | −0.180<br>(0.382) |
| BV |  |  | 1.185***<br>(0.349) |
| SL |  |  | 0.800<br>(0.962) |
| SX |  |  | −0.914<br>(0.722) |
| NRW |  |  | −0.119<br>(0.426) |
| BW |  |  | 0.245<br>(0.466) |
| DAYS | 0.017<br>(0.012) | −0.013<br>(0.012) | −0.011<br>(0.012) |
| ln (r) |  | −1.549***<br>(0.162) | −1.550***<br>(0.168) |
| Constant | −225.876***<br>(54.613) | −62.008<br>(52.268) | −69.584<br>(54.731) |
| Rho | 0.165**<br>(0.064) | 0.042<br>(0.062) | −0.044<br>(0.065) |
| Observations | 412 | 412 | 412 |
| Log Likelihood | −969.231 | −927.965 | −920.308 |
| $\sigma^2$ | 6.435 | 5.293 | 5.100 |
| Akaike Inf. Crit. | 1,964.461 | 1,883.930 | 1,878.615 |
| Wald Test (df = 1) | 6.714*** | 0.449 | 0.454 |
| LR Test (df = 1) | 5.551** | 0.378 | 0.365 |

Note:
\*p<0.1; \*\*p<0.05; \*\*\*p<0.01
Source: own calculation.

**Figure 11:**
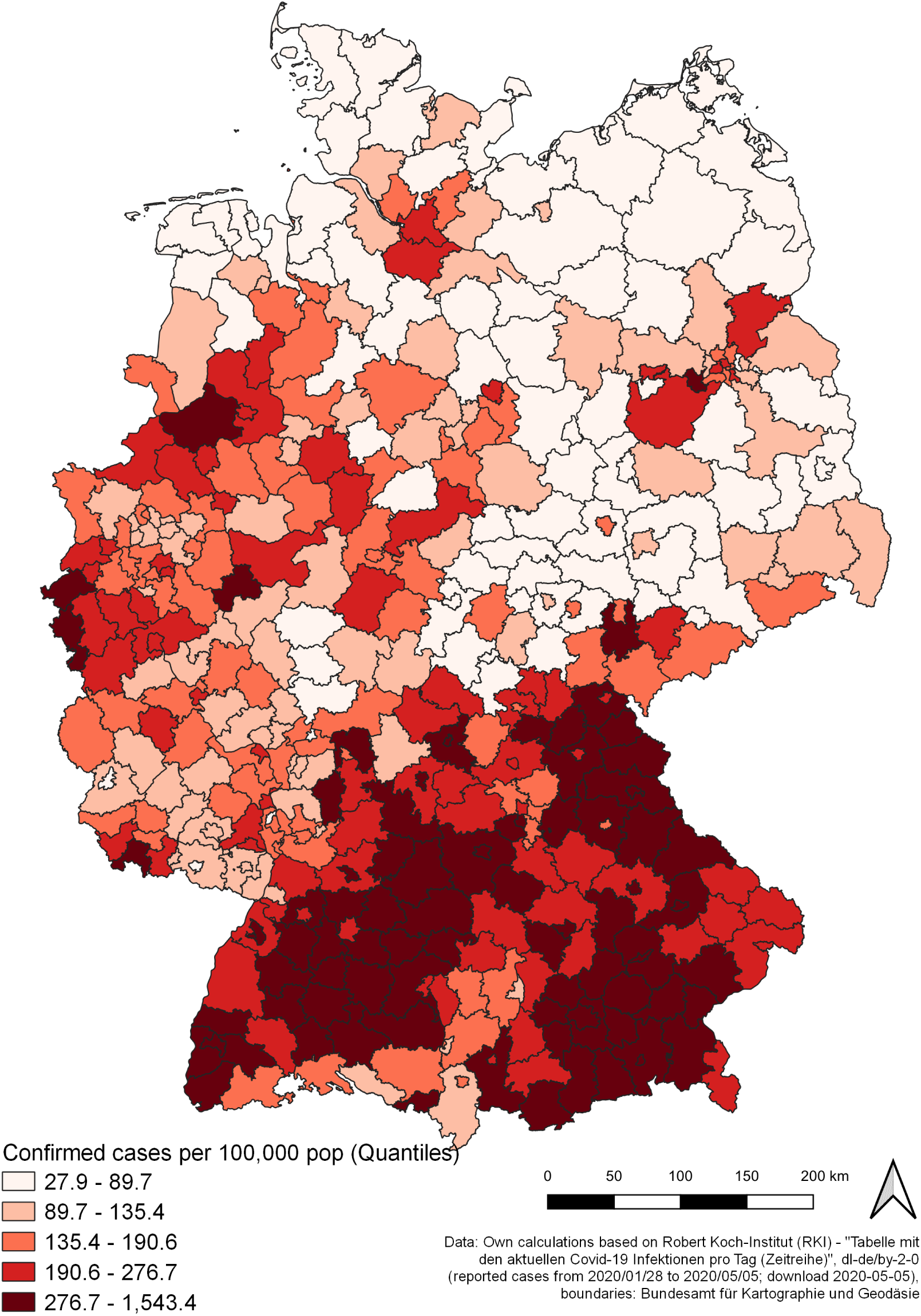
Prevalence by county Source: own illustration.

**Figure 12:**
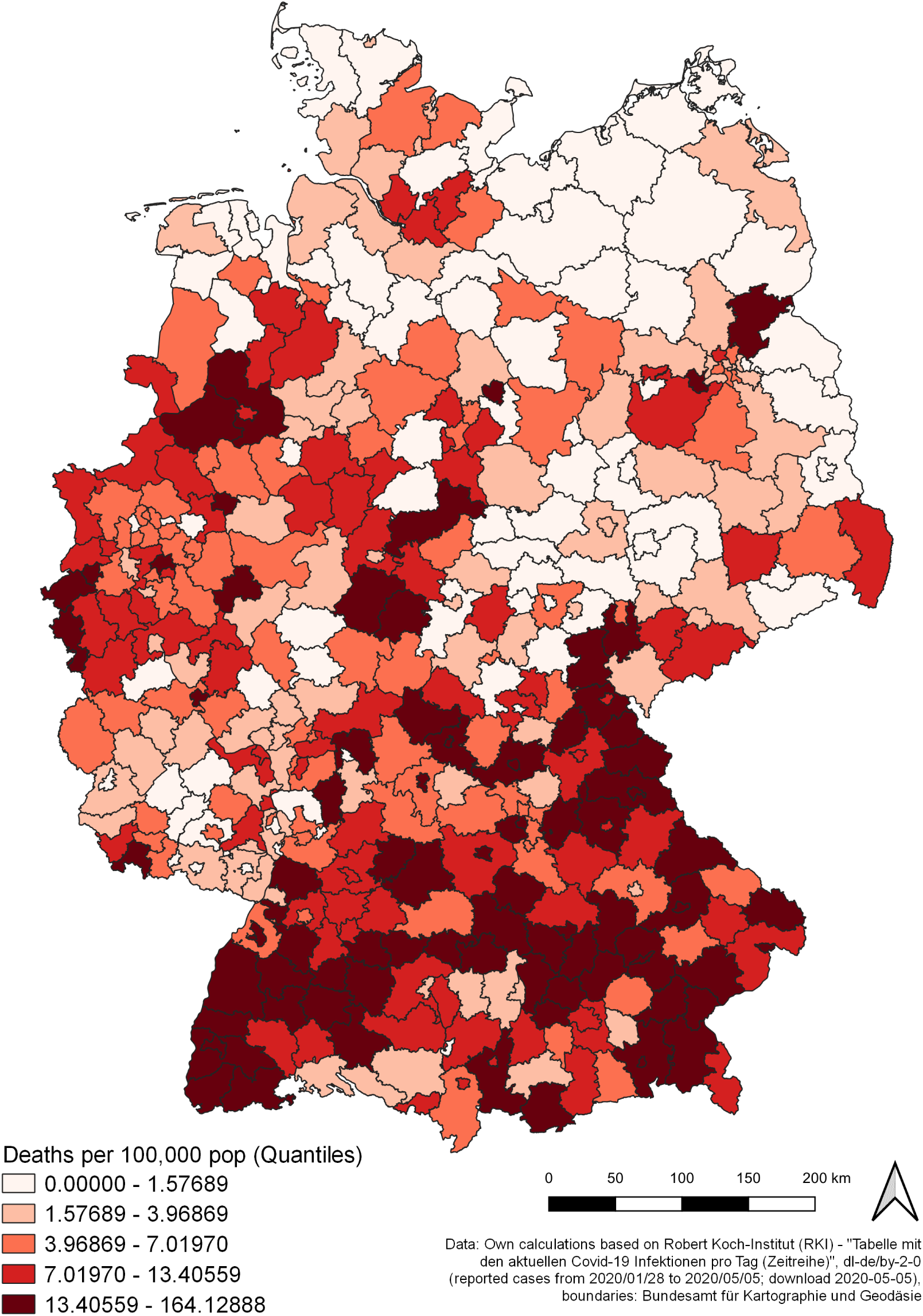
Mortality by county Source: own illustration.

**Figure 13:**
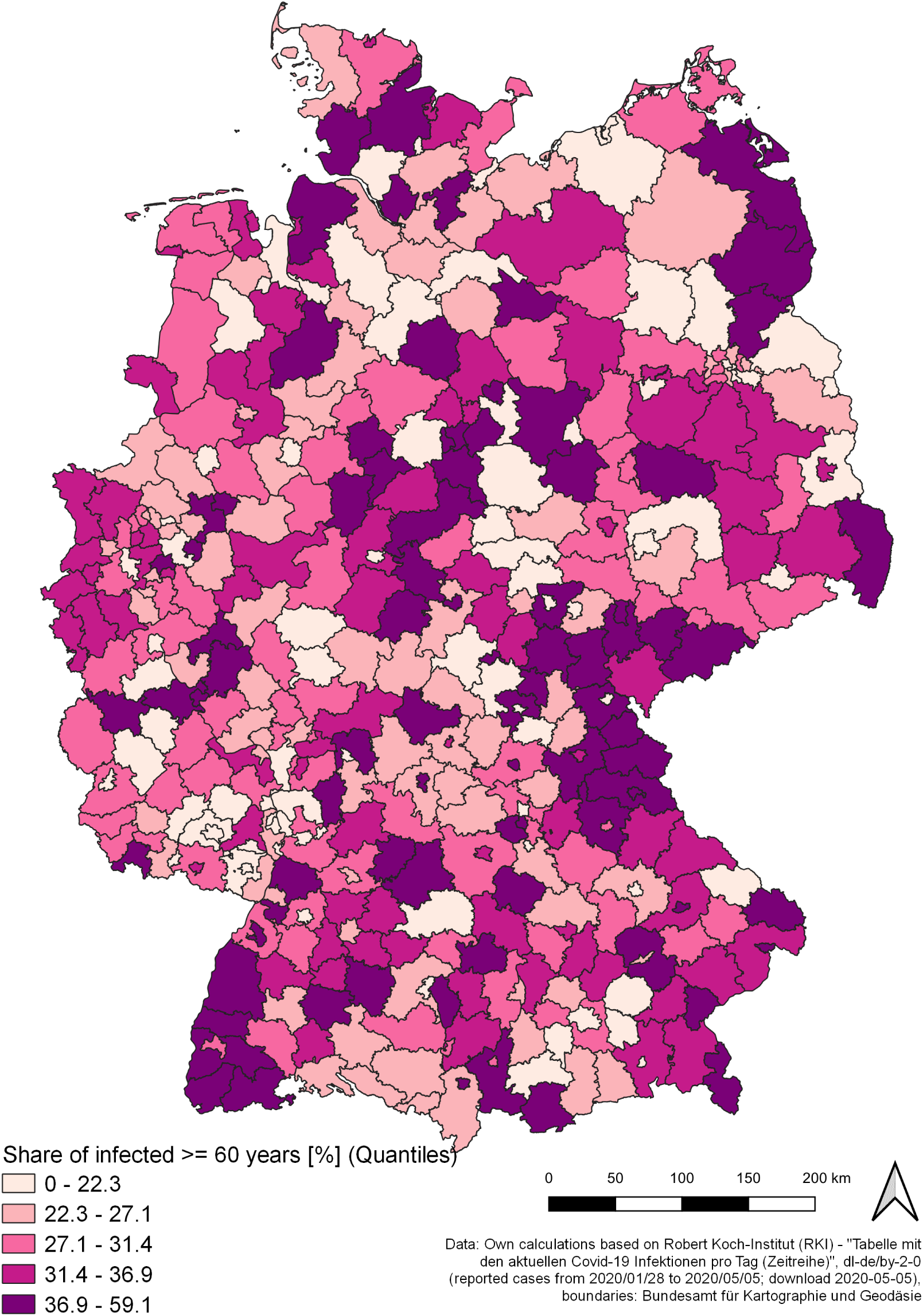
Share of reported infected individuals of age 60 and older by county Source: own illustration.

**Figure 14:**
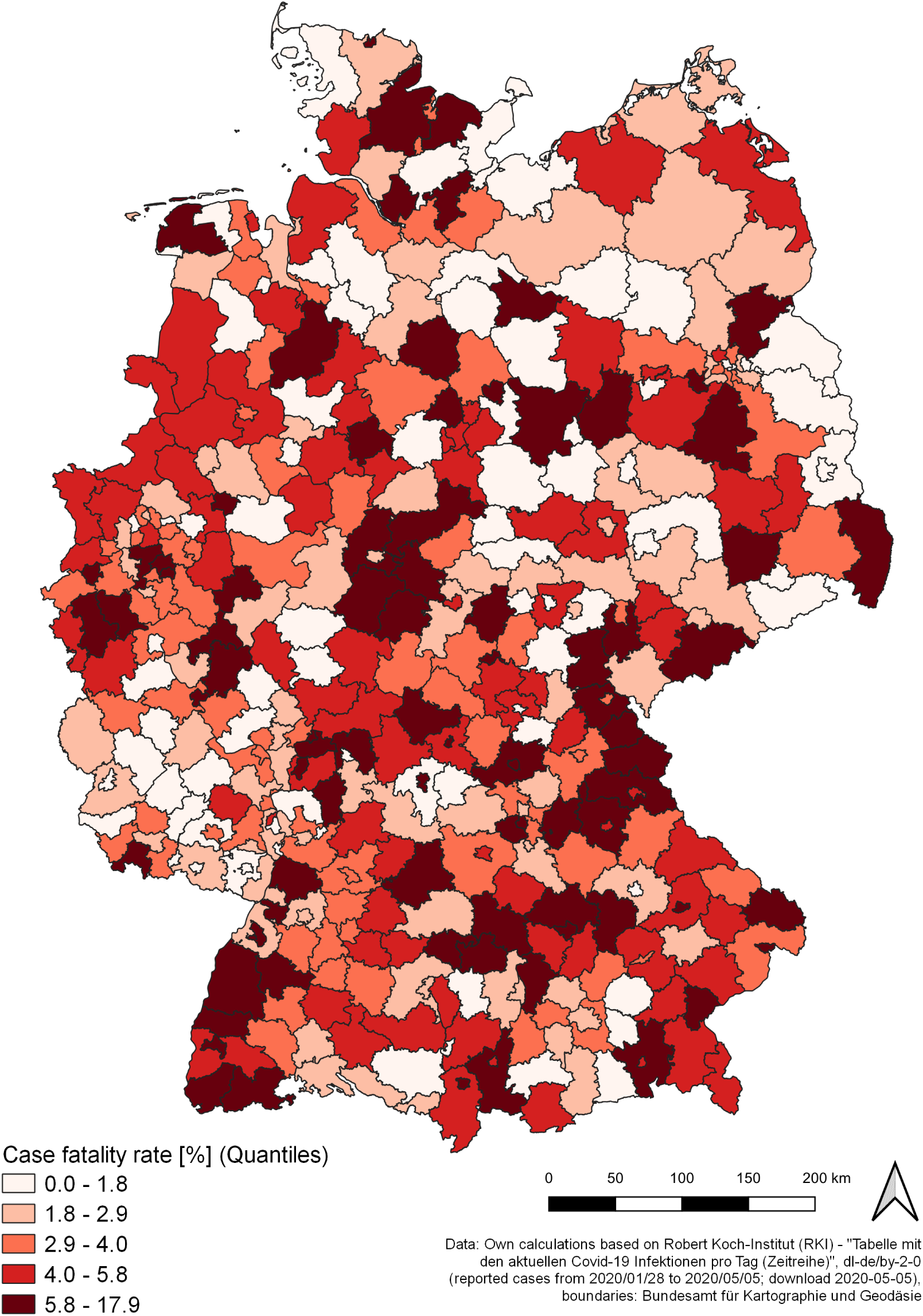
CFR by county Source: own illustration.

In all of the OLS models, no variable exceeded the critical value of *V IF* ≥ 5. For the prediction of the intrinsic growth rate, two model variants were estimated with and without the state dummy variables (table 7). From the aspect of explained variance, the second OLS model provides a better fit (*R*^2^ = 0.731 and *Adj*.*R*^2^ = 0.723, respectively) compared to the first (*R*^2^ = 0.678 and *Adj*.*R*^2^ = 0.672, respectively), thus, the second model is to be preferred for interpretation. Different than expected, population density (*POPDENS*) does not affect growth rate positively. In contrary to the assumptions, a 1% increase of population density decreases the growth rate by 0.1%. Also the demographic indicator (*POPS*65) is correlated with growth in a way which is opposite to the assumed: An increase of 1% in the share of inhabitants of age ≥ 65 increases the growth rate by 1.2 %. As expected, the intensity of commuting (*CMI*) has a significant positive effect on the intrinsic growth rate: An increase of commuting intensity by 1% increases the growth rate by 0.4%. Tourist density (*TOUR*) is only significant in the first model. On average, the intrinsic growth rate is significantly higher in Bavarian counties (dummy variable *BV*), while it is significantly lower in Saxony and North Rhine-Westphalian counties (*SL* and *SX*, respectively). The coefficients for Baden-Wuerttemberg (*BW*) and Saarland (*SL*) are not significant, which means that no significant deviation from the average growth rate is found for counties in these German states. The *EAST* dummy is not significant in both models. Considering the necessary control variables, the current prevalence (*PRV*) and time (*DAY S*) decelerate the growth of infections significantly. The former has a nearly proportional impact: An increase of prevalence equal to 1% decreases the intrinsic growth rate by 1.04%. For each day SARS-CoV-2/COVID-19 is present in the county, the growth speed declines on average by 1.4%. Here, one has to keep in mind that these relationships are reciprocal and represent the mandatory decline of susceptible individuals over time.

For the prediction of mortality (*MRT*), the growth rate (*r*) and the state dummy variables (*BV, SL, SX* and *NRW*) are entered into the model analyis successively, resulting in three models (table 8). When comparing models 1 and 2, adding the growth rate as independent variable increases the explained variance substantially (*Adj*.*R*^2^ = 0.219 and 0.367, respectively). The third model provides the best fit, adjusted for the number of explanatory variables (*Adj*.*R*^2^ = 0.383). No significant influence can be found for the spatial (*POPDENS*), demographic (*POPS*65) and mobility variables (*PI* and *TOUR*) as well as the air pollution variables (*PM* 10 and *NO*2). Life expectancy (*LEXP*) and the dummy for East German counties (*EAST*) are only significant in the first model. However, the share of infected people of age ≥ 60 (*INFS*60) significantly increases the regional mortality: An increase in the share of people of the “risk group” in all infected by 1% increases the mortality by approx. 0.5%. The only significant state-specific effect can be identified with respect to Bavaria: The mortality in Bavarian counties is higher than in the counties belonging to other states. Furthermore, a two-sample t-test reveals that Bavarian counties have a significantly higher share of infected belonging to the risk group 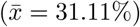 compared to the remaining states 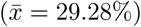, with a difference of 1.83 percentage points (*p* = 0.054). The reciprocal relationship between mortality and growth rate is also significant.

However, as expected, spatial autocorrelation can be detected among the dependent variables. The Moran’s I-statistic for both regional growth rate and mortality (0.49 and 0.16, respectively) is significant (see table 9). By consequence, the OLS estimations might not be robust, which leads to a further exploration of the relationships to be detected in terms of the spatial lag models which incorporate the spatial autocorrelation effect.

With respect to the spatial lag model for regional growth rates (table 10), the *Rho* parameter in both model variants is significant (*Rho* = 0.158 and *Rho* = 0.095, respectively), which indicates a significant spatial lag effect. However, when comparing the second spatial lag model (with *AIC* = 674.96, which is superior to model 1 with *AIC* = 733.37) to the second OLS model (see table 7), there are only negligible differences in the parameter estimates and significance levels, respectively: The same independent variables are found to be significantly correlated with growth rates. They also have the same sign, which indicates the same direction of influence. Intrinsic growth rates on the county level are predicted by population density (approx. −0.9), population share of 65 and older (approx. 1.1), commuting intensity (approx. 0.4), and state-specific dummy variables (Bavaria: approx. 0.5, Saxony: approx. −0.5, North Rhine Westphalia: approx. −0.4) as well as the control variables (Prevalence: approx. −1.0 and time since first infection: approx. −0.01).

The same conclusion must be made with respect to the spatial lag models for mortality (table 11), when comparing them to the OLS models (table 8). The spatial lag effect is only significant in the first model (*Rho* = 0.165) but not in model 2 (*Rho* = 0.042) and 3 (*Rho* = − 0.044). Regional mortality is significantly influenced by the share of infected people of age ≥ 60 (approx. 0.5) and the dummy variable for Bavarian counties (approx. 1.2) as well as correlated with regional growth rates (approx. −1.6). As spatial autocorrelation was also detected for the regional growth rate, which is an independent variable in the mortality models, a further robustness check of the estimations is necessary: Table 12 shows the results for the second and third mortality model in a spatial Durbin model, which incorporates a spatial lag effect for both the dependent variable (*Rho*) and the regional growth rate (*lag ln (r)*). The lag effect of *ln*(*r*) is statistically significant, but the other results remain qualitatively the same (share of infected people of age ≥ 60: approx. 0.5; dummy variable for Bavarian counties: approx. 1).

**Table 12:**
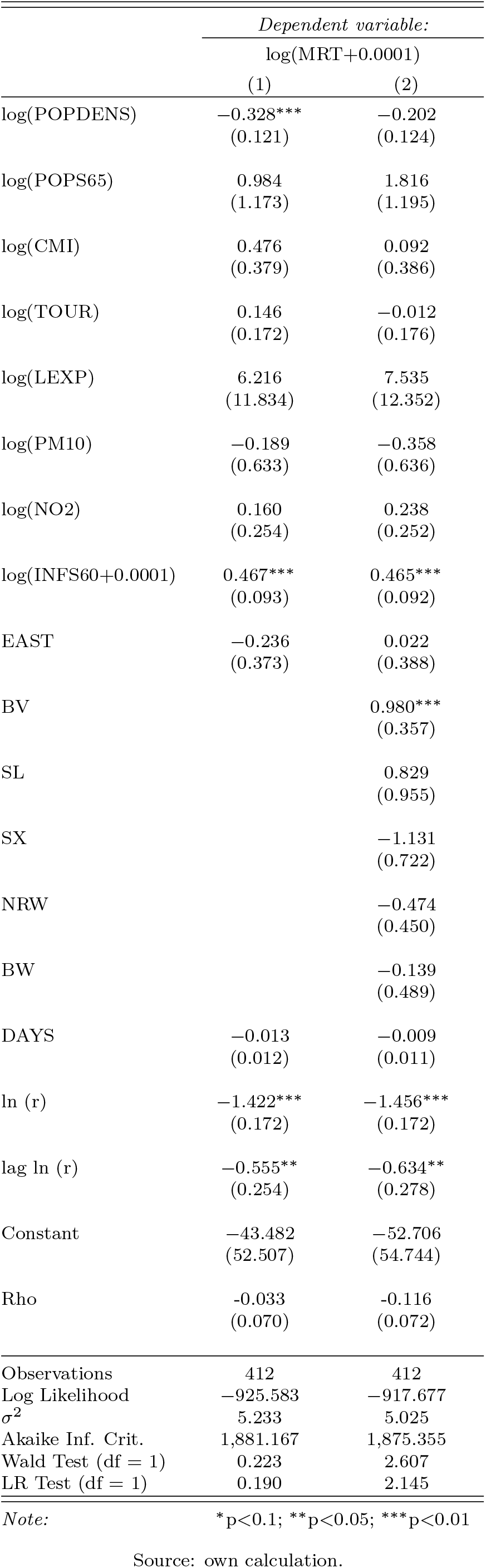
Estimation results for the mortality model (spatial Durbin model)

With respect to the regression models for regional growth and mortality, the results of the OLS estimations were confirmed by those from the models incorporating spatial autocorrelation. Although there is obvious spatial autocorrelation (which can be explained plausibly by interregional transmission of the infectious disease), both OLS and spatial regression models show nearly the same results with respect to strength and direction of correlations. From the spatial statistic point of view, this can be explained with the incorporated independent variables as regional differences in both growth rate and mortality are predicted entirely by the interregional variation in causal factors.

## 4 Discussion

### 4.2 Curve flattening in the context of nonpharmaceutical interventions

Taking a look at the national level, the flattening of the epidemic curve in Germany occurred between three to six days before phase 3 of measures (as defined in this study) came into force. Due to this temporal mismatch, the decline of infections can not be causally linked to the nationwide formal “lockdown” (including forced social distancing and ban of gatherings) of March 23. Note that the results for whole Germany, estimating the inflection point between March 17 and March 20, are rather conservative when compared with the RKI estimations. In the RKI nowcasting study, the peak of onset of symptoms (not infection time, which is not considered in the mentioned study) is found at March 18 and a stabilization of the reproduction number equal to *R* = 1 at March 22 (an der Heiden, Hamouda 2020). Subtracting an average incubation period of five days from these dates, the peak of infections occured around March 13 and the reproduction number stabilizes approximately at March 17. The former RKI study (an der Heiden, Buchholz 2020) estimated the peaks between June and July, depending on the parameters of the scenarios.

The main focus of this analysis is the regional level, which reveals a more differentiated picture. In all German counties the curves of infections clearly flattened within a time period of about six weeks from the first to the last county. On average, it took one month from the first infection to the inflection point of the epidemic curve. However, the regional trend change in infections is not in line with the governmental nonpharmaceutical interventions to contain the virus spread. In nearly two thirds of the German counties which account for two thirds of the German population, the flattening of the infection curve occured before the “lockdown” (measures of phase 3) came into force (March 23). One in eight counties experienced a decline of infections even before the closures of schools, child day care facilities and retail facilities, which is attributed to phase 2 of interventions in this study. Consequently, in a majority of counties, the regional decline of infections can not be attributed to the formal “lockdown”. In a minority of counties, also closures of educational and retail facilities (measures of phase 2) cannot have caused the decline. While keeping in mind that SARS-CoV-2 emerged at different times across the counties, it is at least questionable whether these measures primarily caused the flattening of the infection curve in the other counties. Furthermore, in a minority of counties, the regional trend change occurred several weeks (up to about four weeks) after the nonpharmaceutical interventions came into force. One might argue that there could be a time lag between the date of official enforcement of the regulations and the time they became effective in practice. However, this could only be conceivable for the contact ban but not for the closures of schools and other services as these infrastructures are either closed or not and, thus, can be potential places of virus transmission or not. Moreover, it seems unlikely that an intervention like a contact ban becomes effective only after several weeks and regionally differentiated.

Bringing together these aspects, regional curve flattening seems to have occurred independently from the governmental measures of phase 2 and 3. Instead, regional pandemic growth appears as a function of time, reaching the peak of infection rates with a time lag depending on the date the virus emerged.

The results presented here tend to support the findings in the study by Ben-Israel (2020) that curve flattening in the SARS-CoV-2/COVID-19 pandemic occurs with or without a strict “lockdown”. However, the mentioned study does not provide explicit epidemiological, virological or other kinds of clarifications for this phenomenon, neither the present study does. The further interpretation must be limited to a collection of explanation attempts, which are non-mutually exclusive. Some reasons for the decline of infections relate to other types of interventions both voluntary and mandatory:

- First of all, it must be pointed out that the focus of this study is on regional pandemic growth in the context of the nonpharmaceutical interventions of phase 2 and 3 starting in mid March 2020, especially the “lockdown” from March 23. One has to keep in mind that some interventions against virus spread were already established in the first half of March (phase 1), e.g. the cancellation of large events or “ghost games” in soccer (see table 1). These early measures could have contributed substantially to curve flattening, as the cancellation of events might have prevented people from being infected in the context of so-called *super-spreading events*, which play an enormous role during infectious disease spreads (Al-Tawfiq, Rodriguez-Morales 2020, Stein 2011). Many infections and death cases attributed to COVID-19 in the early phase of the pandemic in Germany can be traced back to super-spreading events in February and early March 2020, such as in Heinsberg or Tirschenreuth county (Tagesspiegel 2020b). Also, the domestic quarantine of infected persons (which is the default procedure in the case of infectious diseases) might have reduced new infections. In Heinsberg county, about 1,000 people were in domestic quarantine at the end of February 2020 (Tagesschau.de 2020c), which could explain the early curve flattening in this Corona “hotspot”.
- Also media reports from China or Italy as well as appeals and recommendations from the government could have influenced people to change their behavior on a *voluntary* basis already in the first half of March 2020 (or even earlier), e.g. with respect to physical distancing, thorough and frequent hand washing, coughing and sneezing in the arm fold, or reducing mobility in general. Unfortunately, there is no explicit indicator of changes in the individual behavior. However, some other findings give a hint towards voluntary behavorial changes: Several surveys show a high degree of public awareness in Germany (and other countries) towards the SARS-CoV-2/COVID-19 threat already in February and the first half of March 2020 (Ipsos 2020, YouGov 2020). This increasing awareness might be reflected by more caution in daily life: The RKI has documented an “abrupt” decline of *other* infectious respiratory diseases with shorter incubation periods (such as influenza) in Germany since the 10th calendar week (March 2 to 8, 2020). This decline is regarded as “extremely unusual” (Buchholz et al. 2020). This reduction might be attributed to voluntary cautious behavior in the context of the public discussion towards SARS-CoV-2, as this decrease started before any public health intervention came into force (except for the quarantines of SARS-CoV-2-infected persons, see table 1). Furthermore, the analysis of mobility patterns shows a decline of mobility in Germany, starting already in the first half of March 2020. Additionally, a strong correlation between (aggregated) mobility and the acceptance of social restrictions (obtained by surveys) was found: The higher the agreement with the statement “I think the current measures are too strict”, the higher the increase in mobility (Covid-19 Mobility Project 2020a,b). All these phenomenons suggest voluntary behavior changes within the (German) population, which reduce the transmission of infectious diseases and preceded the “lockdown” by several weeks. Another indicator for an increased awareness in the (German) population - although not intended or desired - is the enormous tendency of hoarding groceries, which started in the second half of February 2020 (Rheinische Post online 2020).
- Additionally, one has to keep in mind the seasonal cycle of respiratory viral diseases: Influenza viruses and most cold viruses (including those from the family of Coronaviridae) mainly occur during the winter months due to changes in environmental parameters (e.g., temperature and humidity) and human behavior (more or less activities outside, whilst the risk of infection is, all other things being equal, less outside) (Moriyama et al. 2020). Several virologists expressed cautious optimism towards the sensitivity of the SARS-CoV-2 virus to increasing temperature and ultraviolet radiation (Focus 2020). Model-based analyses from biogeography show that temperate warm and cold climates facilitate the virus spread, while arid and tropical climates are less favorable (Araujo, Naimi 2020). By consequence, there might have also been a decline of SARS-CoV-2 infections due to weather changes in early spring (mid-March).

Furthermore, there is another possible reason for curve flattening with or without a “lockdown”, which is of epidemiological nature and related to the transmission process of SARS-CoV-2/COVID-19 in the context of immunization. Note that “immunity” may have different causes (e.g., antibodies due to previous infections, vaccination, immunological memory) and does not necessarily prevent individuals from being infected (in terms of an invasion of an individual’s body) but leads to an effective response of the immune system and prevents the emergence of (severe) symptoms (Mak, Saunders 2006). At the beginning of the pandemic, two assumptions towards the role of immunization were stated: 1) Nobody is immune, which means that all individuals of the population belong to the group of susceptibles, 2) Without any interventions (e.g., vaccine, nonpharmaceutical interventions), *herd immunity* - a share of a population is immune, which provides protection to those who are not immune, causing the pandemic to slow down and stop - is achieved when about 70% of the population was infected and recovered (D’Souza, Dowdy 2020). This percentage share is commonly known as *herd immunity threshold* (*HIT*) and is, in its basic form, calculated based on the infection’s basic reproduction number, *R*_0_: *HIT* = 1 − 1*/R*_0_ (Fine et al. 2011). Also early modeling studies focusing the effect of nonpharmaceutical interventions are based on these (or similar) assumptions (an der Heiden, Buchholz 2020, Ferguson et al. 2020). However, there are some issues regarding the *HIT* for SARS-CoV-2/COVID-19 which need to be considered:

- In epidemiology, it is well known that disease transmission is mostly concentrated on a minor part of individuals causing a large majority of secondary infections: “In what became known as the 20/80 rule, a concept documented by observational and modeling studies and having profound implications for infection control, 20% of the individuals within any given population are thought to contribute at least 80% to the transmission potential of a pathogen, and many host-pathogen interactions were found to follow this empirical rule” (Stein 2011). Gomes et al. (2020) incorporate inter-individual variation in susceptibility and exposure to a SARS-CoV-2 infection into an epidemiological model (SEIR [susceptible-exposed-infectious-recovered] model). Depending on the assumptions on this *overdispersion*, the *HIT* of SARS-CoV-2 reduces to 10-20%. Thus, the achievement of herd immunity would require a considerably lower number of SARS-CoV-2 infections.
- Although the SARS-CoV-2 virus is highly infective, the “Heinsberg study” by Streeck et al. (2020) found a relatively low secondary infection risk (*secondary attack rate*, SAR). Infected persons did not even infect other household members in the majority of cases. The authors conjecture that this could be due to a present immunity (T helper cell immunity) not detected as positive in the test procedure. This kind of immunity is not to be confused with (temporal or everlasting) immunity due to antibodies against a specific virus but may be regarded as a functional immune memory. In a current virological study by Braun et al. (2020), 34% of test persons who have never been infected with SARS-CoV-2 had relevant T helper cells because of earlier infections with other harmless Coronaviruses causing common colds. In a study by Grifoni et al. (2020), SARS-CoV-2-reactive T cells were detected in even 40%–60% of unexposed individuals. If this explanation proves correct, the absolute number of susceptible individuals would have been substantially lower already at the beginning of the pandemy. Other Coronaviruses are responsible for about 10-15% of seasonal “common colds” (Padberg, Bauer 2006). Cross protection due to related virus strains has also been determined e.g. with respect to influenza viruses (Broberg et al. 2011).
- Considering the aforementioned aspects, we have to keep in mind that all data related to infections used here underestimate the real amount of infected individuals in Germany as well as in nearly all countries in which the Coronavirus emerged. Typically, at the beginning of the pandemic, only suspected cases with COVID-19 symptoms were tested, leading to a heavy underestimation of infected people without symptoms (see section 2.2). Several recent studies have tried to estimate the real prevalence of the virus and/or the infection fatality rate (IFR), including all infected cases rather than the confirmed (see table 13). Estimated rates of unreported cases (estimate PRV/reported PRV) lie between 5 (Gangelt, Germany) and 50-85 (Santa Clara County, USA). Obviously, when estimated CFR values exceed the estimated IFR values by ten times or more, there must be a large amount of unreported cases and the total number of infected individuals must be considerably higher than reported, respectively. The logical consequence is that there is a hidden decrease of the absolute number of susceptible individuals because of many infected persons without symptoms not knowing that they have been infected (and probably immunized) in the past. These individuals were not tested for the *pathogen* (SARS-CoV-2 virus) because they did not suffer from the *disease* (COVID-19). Quantifying the “dark figure” of SARS-CoV-2 infections by using representative sample-based tests on current infection as well as seroprevalence will be a challenge in the near future.

Of course, the present empirical results cannot prove or disprove the presence or absence of (herd) immunity. However, the number of infected individuals is obviously quite higher than reported (see table 13), whilst the number of susceptibles could have been considerably smaller than expected already at the beginning of the pandemic. If a SARS-CoV-2 infection leads to (lifelong or temporal) immunity (which is not yet clarified), the current level of immunity must be higher as well. Similar results were found in the UK: Stedman et al. (2020) also find decreasing infection rates (*R*_*ADIR*_) related to (reported and unreported) prevalence on the regional level (Upper Tier Local Authority areas) and conclude that “the only factor that could be related to the *R*_*ADIR*_ in this analysis was the historic number of confirmed number infection/,000 population suggesting that some of the reduction in reported cases is due to the build-up of immunity due to larger numbers of historic cases in the population”.

**Table 13:** Studys on unreported cases and/or IFR of SARS-CoV-2/COVID-19

| Study | Study area | n | Time of data collection | Est. PRV [%] (CI95) | Est.asymp-tomatic cases [%] | Est. PRV/ reported PRV | Est. IFR [%] (CI95) |
| --- | --- | --- | --- | --- | --- | --- | --- |
| Bendavid et al. (2020) | Santa Clara county (USA) | 3,324 | 04/2020 | 2.8<br>(2.2, 3.4) | NA | 50-85 | NA |
| Bennett, Steyvers (2020)<br>- re-analysis of<br>Bendavid et al. (2020) | Santa Clara county (USA) | 3,324 | 04/2020 | 0.27-3.21 | NA | 5-65 | NA |
| Gudbjartsson et al. (2020) | Iceland |  |  |  |  |  |  |
| Targeted testing 1 |  | 177 | 01-03/2020 | 9.2 | 13.6 | NA | NA |
| Population screening 1 |  | 10,797 | 03/2020 | 0.8 | 41.4 | NA | NA |
| Targeted testing 2 |  | 7,275 | 03/2020 | 14.4 | 5.7 | NA | NA |
| Population screening 2 (Random sample) |  | 2,283 | 04/2020 | 0.6 | 53.8 | NA | NA |
| LAPH (2020) | Los Angeles county (USA) |  | 04/2020 | 4.1<br>(2.8, 5.6) | NA | 28-55 | NA |
| Lavezzo et al. (2020)<sup>1</sup> | Vo (Italy) |  |  |  |  |  |  |
| First survey |  | 2,812 | 02/2020 | 2.6<sup>2</sup><br>(2.1, 3.3) | 41.1 | NA | NA |
| Second survey |  | 2,343 | 03/2020 | 1.2<sup>2</sup><br>(0.8, 11.8) | 44.8 | NA | NA |
| Nishiura et al. (2020)<sup>1</sup> | Wuhan (China)<sup>3</sup> | 565 | 02/2020 | 1.4 | 62.5 | 5-20 | 0.3-0.6 |
| Russell et al. (2020)<sup>1</sup> | Diamond Princess cruise ship | 3,711 | 02/2020 | 17 | 51.4 | NA | 1.3<br>(0.38, 3.6) |
| Shakiba et al. (2020) | Guilan province (Iran) | 551 | 04/2020 | 33<sup>4</sup><br>(28, 39) | 18 | NA | 0.08-0.12 |
| Streeck et al. (2020) | Gangelt (Germany) | 919 | 03/2020 | 15.5<br>(12.3, 19.0) | 22.2 | 5 | 0.36<br>(0.29, 0.45) |

Source: own compilation.
Notes: <sup>1</sup>full survey or nearly full survey, <sup>2</sup>only active infections (not including seroprevalence), <sup>3</sup>Japanese citizens evacuated from Wuhan (China), <sup>4</sup>adjusted for test performance.

However, we have to keep in mind that even if herd immunity was achieved, this does not mean that *no* new infections occur. Furthermore, herd immunity implies a closed population, where there are too many immunized individuals to infect the remaining susceptibles. In reality, there are migratory and mobility flows between regions and nations (e.g., work-related commuting, tourism) and, thus, new infections may occur due to transmissions driven by spatial interactions. More precisely, a susceptible individual living in a given region with herd immunity might get infected when traveling to another region. Finally, there is a difference between *infection* and *disease*, whilst the infectiousness of asymptomatically infected individuals is not yet clarified, although they are regarded as much less likely to transmit the virus than infected with symptoms (World Health Organization 2020a).

### 4.2 Determinants of regional growth and mortality

Two regression models were estimated, including intrinsic growth rates (indicating the speed of pandemic growth) and mortality (indicating the severity of the disease) as dependent variables. For both variables, spatial autocorrelation was detected, which can be explained comprehensively by virus transmission across borders of nearby counties. However, both the OLS and the spatial regression models give qualitatively the same results with respect to the correlations.

With respect to the determinants of growth speed, two explanatory variables have a significant effect which is opposite to the expected: A slower disease spread is not due to lower population density. In contrast, intrinsic growth rates decrease with higher density values on the level of German counties. This could be explained with the validity of this indicator as it is questionable whether population density is a sufficient proxy for the amount and intensity of physical contacts between individuals. Although living in larger and densely populated municipalities (such as large cities), there is no empirical evidence that inhabitants automatically have more social interactions than in rural areas (Mitterer 2013, Petermann 2001). There is also no dampening effect of virus spread by an older population on the county level. Instead, growth rates increase with the share of inhabitants of 65 years and older, although this age category covers the retired population. This result might be due to a bias in testing for SARS-CoV-2 infections: Most tests in the past were conducted on people with COVID-19 symptoms. As older people are more likely to have a severe course of the disease, these age groups are obviously overrepresented in the confirmed cases (RKI 2020a). Differing from the expectations, no isolated effect of East German counties was found.

However, regional growth rates of infections are increased by inter-regional mobility, especially with respect to work-related commuting. This result is quite plausible and supports previous findings in the literature on empirical and model-based approaches towards infectious disease spreads in the past (Charaudeau et al. 2014, Dalziel et al. 2014, Findlater, Bogoch 2018).

Taking a look at state-specific effects, the results show a significant higher average growth rate in Bavarian counties and lower growth rates in Saxony and North Rhine Westphalia, while Saarland and Baden-Wuerttemberg counties did not deviate from the all-over average. Thus, no dampening effect on pandemic growth can be confirmed for German states with additional curfews for the containment of the virus spread (Bavaria, Saarland, Saxony). The growth rate in Bavaria is even considerably above the average, although the time since the virus emerged and the current prevalence were included into the models as control variables.

With respect to the determinants of mortality, few clear statements can be made. SARS-CoV-2/COVID-19 mortality on the county level is not significantly influenced by demographic, spatial or mobility factors. However, these variables explain the regional growth rate, which was added to the mortality model as control variable (and is significant, as expected). There is no specific “East Germany effect” as well.

Considering previous studies on the influence of air pollution on COVID-19 severity (Ogen 2020, Wu et al. 2020), it was expected that particulate matter and *NO*_2_ concentration would have an influence on mortality. Although it is plausible to assume that air pollution increases fatality rates of respiratory diseases, this hypothesis was not confirmed in the present study, which may result from an obvious data problem: The annual mean values of daily pollution was obtained on the level of monitoring stations, which are not evenly distributed across the counties and measure the air pollutant level at a specific point (e.g. traffic crossroad). It is unlikely that these obtained values are representative for the whole county. Thus, the validity of this indicator is questionable. Unfortunately, county-based data towards air pollution is not available nationwide.

Obviously, the variance of regional mortality reflects the regional variance of infected individuals belonging to the “risk group”, defined as people of 60 years and above. Although no regional data is available for cases and deaths in retirement homes, a large share should be attributed to these facilities. Nationwide, people accommodated in facilities for the care of elderly make up at least 2,473 of 6,831 deaths (36.20 %) as of May 5, 2020 (RKI 2020a). The share of residents of retirement homes in all COVID-19-related deaths is equal to 51% in France and 33% in Denmark, ranging internationally from 11% in Singapore to 62% in Canada (Comas-Herrera et al. 2020). The relevance of retirement homes in Germany can be underlined with examples based on information available in local media which depicts the regional situation:

- In the city of Wolfsburg (Lower Saxony), the current mortality (MRT) is equal to 41.08 deaths per 100,000 inhabitants, while the current case fatality rate (CFR) is the highest in all German counties (17.89 %). Both values are calculated from the data used here (of date May 5, 2020). As of May 11, there have been 51 deaths attributed to COVID-19 in Wolfsburg, with 44 of these deaths (86.27 %) stemming from residents of one retirement home (Wolfsburger Nachrichten 2020).
- In the Hessian Odenwaldkreis with MRT = 54.75 and CFR = 14.60 %, 29 people who tested positive to SARS-CoV-2/COVID-19 died until April 14, 2020, 21 of them (72.41 %) were living in retirements homes in this county (Echo online 2020).
- In the city of Würzburg (Bavaria) with MRT = 38.32 and CFR = 10.75 %, there have been 44 COVID-19 positive deceased in two retirement homes until April 24, 2020, leading to investigations by the public prosecution authorities (BR24 2020). Up to April 23, 2020, in the whole administrative district Unterfranken, 64 % of all people who died from or with Corona were residents of retirement homes for elderly people (Mainpost 2020).

With respect to state-specific effects, there is a clear significant impact regarding Bavaria: Although the measures in Bavaria, based on the Austrian model, were probably the strictest of all German states, both regional growth rates and mortality are significantly *higher* than in the other states. This effect is isolated, as other effects (time, population density etc.) were controlled. In addition, the share of individuals belonging to the “risk group” is slightly higher in Bavaria. In the other states with curfews, Saarland and Saxony, no significant impact of this additional intervention was found, especially with respect to mortality which does not differ significantly from other states.

## 5 Conclusions and limitations

In the present study, regional SARS-CoV-2/COVID-19 growth was analyzed as an empirical phenomenon from a spatiotemporal perspective. Using infection dates estimated from reported cases, logistic growth models were estimated for the disease spread at the level of German counties as well as at the national level. The resulting intrinsic growth rates vary across the 412 German counties. The inflection points of the epidemic curves were contrasted to the dates where nonpharmaceutical interventions against the disease spread came into force. As a result, whole Germany as well as the majority of German counties have experienced a decline of the infection rate - which means a flattening of the infection curve - before the main social-related measures (contact ban, ban of gatherings and closure of “nonessential” services) were established. In a minority of counties, curve flattening even occured before schools and child day care facilities were closed. In contrast, some regional trend changes took place several weeks after the measures came into force. *Due to this temporal mismatch, we have to conclude that the decline of infections can not be causally linked to the “lockdown” of March 23. Moreover, also the impact of school and child care infrastructure closures on the pandemic spread remains questionable*.

However, this does not mean that the disease spread slows down automatically. Four possible reasons have been identified for curve flattening independent from school closures and the “lockdown”, with the first two relating to other types of (state-run and voluntary) measures which could reduce the transmission of an infectious disease: 1) Positive effects of *previous governmental nonpharmaceutical interventions* (especially the cancellation of large-scale events), 2) *voluntary behavior changes* (e.g., with respect to physical distancing and hygiene), 3) *seasonality* of the virus, and 4) a rising but undiscovered level of *immunity* within the population. However, whether these determinants may have contributed to the decline of infections, is outside the scope of the model-based analysis.

The determinants of regional intrinsic growth rates (as an indicator for the speed of pandemic spread) and mortality (as an indicator of the disease’s severity) were explored using regression models. Among other things, regional pandemic growth is found to be driven by inter-regional mobility. Mortality on the county level obviously depends on the level of infected individuals belonging to the “risk group” (people of age 60 or older). This share is considerably influenced by SARS-CoV-2/COVID-19 outbreaks in retirement homes for the eldery, which have occurred in many German counties. Obviously, neither strict measures in Germany nor other countries were able to prevent these location-specific outbreaks. *By consequence, it must be concluded that the severity of SARS-CoV-2/COVID-19 depends on the local/regional ability to protect the “risk group”, especially older people in care facilities. This is the more important as virus transmission in care homes is nearly independent from the nonpharmaceutical interventions concerning e*.*g. schools, commercial services, and private residences*. Three German states (Bavaria, Saarland, Saxony) established curfews additional to the nationwide interventions. *We must conclude that these state-specific curfews did not contribute to a more positive outcome with respect to growth speed and mortality*.

On the one hand, these findings pose the question as to whether contact bans and curfews are appropriate measures for containing the virus spread, especially when weighing the effects against the social and economic consequences as well as the curtailment of civil rights. One the other hand, when looking at regional mortality and case fatality rate, the protection of “risk groups”, especially older people in retirement homes, is obviously of moderate success.

From the methodological point of view, two further conclusions must be stated: Nonpharmaceutical interventions aim at the reduction of new infections, thus, their impact must be assessed regarding temporal coincidences with new infections. *Regardless of the modeling approach used for the analysis of pandemic spread, any analysis concerning the effectiveness of nonpharmaceutical interventions must be based on realistic infection dates rather than reporting dates of infected persons. An over- or underestimation of the time between infection and report - in particular, the reporting delay - might lead to senseless conclusions towards the influence of specific measures*. Estimating the true infection dates from reported cases in official statistics is the biggest methodological challenge in this context. Moreover, the present study reveals the importance of a spatial perspective on pandemic spread: *Spatially varying growth rates and severity measures show that the spread of an infectious disease is to be regarded as a spatiotemporal phenomenon. Thus, further studies should address regional differences of epidemiological variables with respect to transmission*.

However, despite these conclusions, the study is faced with two important limitations:

- One has to keep in mind that previous model-based simulation studies which prove the effectiveness of nonpharmaceutical interventions already make *a priori* assumptions on the impact of these measures: In particular, the *input* parameters of the epidemiological models (such as the intensity of physical contacts between individuals) are set in a way that interventions (such as school closures or social distancing) reduce the transmission of the virus in any case and, thus, the simulation *output* shows a decline of infections subsequent to these interventions (an der Heiden, Buchholz 2020, Ferguson et al. 2020). This type of modeling approach (and the corresponding results) might be regarded either as a “causal model” or a tautology. In contrast, the modeling approach used here is of purely empirical nature, only incorporating time series of infections. By consequence, the results are not causal but correlative with respect to the presence or absence of temporal coincidences. It can be shown that curve flattening does not coincide with the focused interventions but occurred after previous interventions and might be due to several other causes. However, the actual reasons *why* the infections declined cannot be deduced from modeling results but must be explored based on interpretations of several empirical hints. Furthermore, as the focus is on inflection points and trend changes, respectively, the present empirical analysis cannot rule out *additional* impacts of the German “lockdown”, e.g., in terms of a stabilization effect.
- Finally, it is necessary to take a look at the informative value of the data on reported cases of SARS-CoV-2/COVID-19 used here. While several statistical uncertainties have been addressed by estimating the dates of infection in the present study, the method of data collection has not been made a subject of discussion. The confirmed cases of infections reported from regional health departments to the RKI result from SARS-CoV-2 tests conducted in the case of specific symptoms. When aggregating the reported cases to time series and analyzing their temporal evolvement, it is implictly assumed that the amount of tests remains the same over time. However, the number of tests was increased enormously during the pandemic - which is to be welcomed from the point of view of public health. From a statistic perspective, it might cause a bias because an increase in testing *must* result in an increase of reported infections, as a larger share of infections is revealed, all other things being equal. In his statistical study, Kuhbandner (2020) argues that the detected SARS-CoV-2 pandemic growth is mainly due to increased testing, leading to the conclusion that “the scenario of a pandemic spread of the Coronavirus in based on a statistical fallacy”. To confirm or deny this conclusion is not subject of the present study. However, taking a look at the conducted tests per calendar week (see figure 15) reveals weekly differences in the amount of tests. From calendar week 11 to 12, there has been an increase of conducted tests from 127,457 (5.9 % positive) to 348,619 (6.8 % positive), which means a raise by factor 2.7. The maximum of tests was conducted in CW 14 (408,348 with 9.0 % positive results), decreasing beyond that time, rising again in CW 17. The absolute number of positive tests is reflected plausibly in the number of reported cases (green line), as the confirmed cases result from the tests. The most estimated infections occurred in CW 11 and 12, showing again the delay between infection and case confirmation. Apart from the fact that excessive testing is probably the best strategy to control the spread of a virus, the resulting statistical data may suffer from underestimation and overestimation, dependent on which time period is regarded.

**Figure 15:**
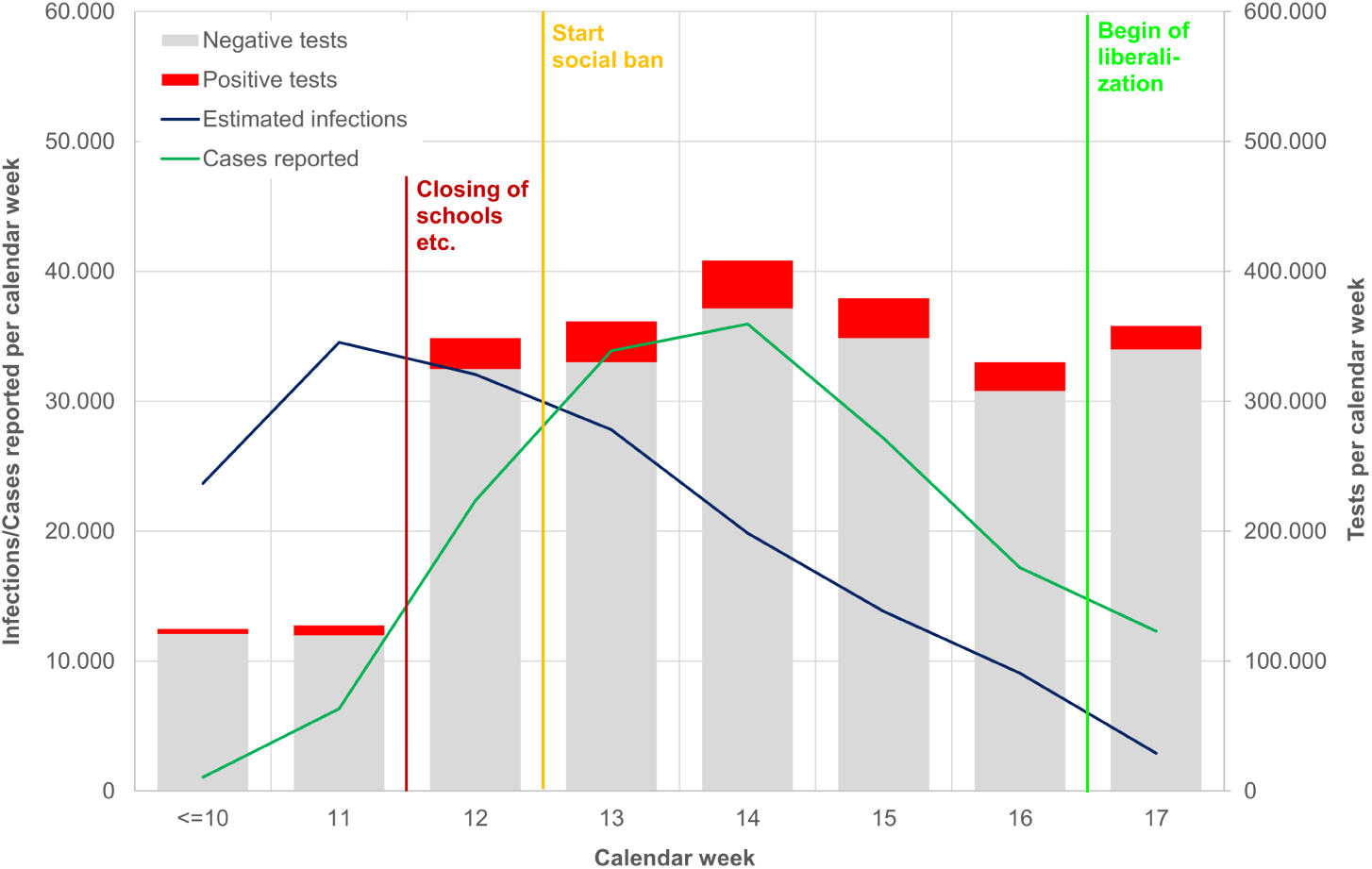
Estimated infections, reported cases and conducted tests by calendar week Source: own illustration. Data source: own calculations based on RKI (2020b,c)

## Data Availability

No patient data etc. was used

## Notes

### Competing Interest Statement

The authors have declared no competing interest.

### Funding Statement

No external funding was received.

### Author Declarations

No patient data etc. was used

